# Spirometric classifications of COPD severity as predictive markers for clinical outcomes: the HUNT Study

**DOI:** 10.1101/2020.11.03.20221432

**Authors:** Laxmi Bhatta, Linda Leivseth, Xiao-Mei Mai, Anne Hildur Henriksen, David Carslake, Yue Chen, Pablo Martinez-Camblor, Arnulf Langhammer, Ben Michael Brumpton

## Abstract

**Rationale:** GOLD grades based on percent-predicted FEV_1_ poorly predicts mortality. Studies have recommended alternative expressions of FEV_1_ for the classification of COPD severity and they warrant investigation.

**Objective:** To compare the predictive abilities of ppFEV_1_ (ppFEV_1_ quartiles, GOLD grades, ATS/ERS grades), FEV_1_ z-score (FEV_1_ z-score quartiles, FEV_1_ z-score grades), FEV_1_.Ht^-2^ (FEV_1_.Ht^-2^ quartiles, FEV_1_.Ht^-2^ grades), FEV_1_.Ht^-3^ (FEV_1_.Ht^-3^ quartiles), and FEV_1_Q (FEV_1_Q quartiles) to predict clinical outcomes.

**Methods:** People aged ≥40 years with COPD (n=890) who participated in the HUNT Study (1995-1997) were followed for 5 years (short-term) and up to 20.4 years (long-term). Survival analysis and time-dependent area under curve (AUC) were used to compare the predictive abilities. A regression tree approach was applied to obtain optimal cut-offs of different expressions of FEV_1_. The UK Biobank (n=6495) was used as a replication cohort with a 5-year follow-up.

**Results:** As a continuous variable, FEV_1_Q had the highest AUCs for all-cause mortality (short-term 70.2, long-term 68.3), respiratory mortality (short-term 68.4, long-term 67.7), cardiovascular mortality (short-term 63.1, long-term 62.3), COPD hospitalization (short-term 71.3, long-term 70.9), and pneumonia hospitalization (short-term 67.8, long-term 66.6), followed by FEV_1_.Ht^-2^ or FEV_1_.Ht^-3^. Generally, similar results were observed for FEV_1_Q quartiles. The optimal cut-offs of FEV_1_Q had higher AUCs compared to GOLD grades for predicting short-term and long-term clinical outcomes. Similar results were found in UK Biobank.

**Conclusions:** FEV_1_Q best predicted the clinical outcomes and could improve the classification of COPD severity.

## INTRODUCTION

The classification of COPD severity is important in guiding therapy and prognosis(1). The Global Initiative for Chronic Obstructive Lung Disease (GOLD) has recommended GOLD grades(1) and the American Thoracic Society (ATS)/ European Respiratory Society (ERS) has recommended ATS/ERS grades(2) based on post-bronchodilator (BD) percent-predicted forced expiratory volume in first second (ppFEV_1_), which is widely used in respiratory medicine. However, ppFEV_1_ has been criticized due to its susceptibility to physiological variation(3-5). Studies have shown that ppFEV_1_ is a poor predictor of mortality(6-8). Accordingly, GOLD grades, which are based on ppFEV_1_, poorly predict mortality(6, 8). Hence, studies have recommended alternative expressions of FEV_1_ and classification of COPD severity(3, 4, 6, 9-12).

FEV_1_ z-score has been recommended as it not susceptible to variation in people’s age, sex, height, and race(3, 4, 13). Other expressions of FEV_1_ that have been recommended included FEV_1_ standardized by squared height (FEV_1_.Ht^-2^)(6, 12) and cubic height (FEV_1_.Ht^-3^)(9, 11, 14), which account for size differences and indirectly for some sex differences. Miller et al.(9) recommended the FEV_1_ quotient (FEV_1_Q), where FEV_1_ is standardized by sex-specific lowest percentile of FEV_1_ distribution that takes account of sex and some size differences in lung function(9). Some studies have investigated these FEV_1_ expressions and/or their respective classifications of COPD severity(5-7, 9, 10, 14-17). Among them, only Huang et al.(5) and Hegendorfer et al.(15) have compared the FEV_1_ expressions mentioned above to predict mortality and exacerbation(5) or all-cause hospitalizations(15). However, Huang et al.(5) had a relatively small sample of people with COPD (n=296) and the study by Hegendorfer et al.(15) included 501 people, among whom only 70 had asthma/COPD. No study has compared the predictive abilities of a broad range of FEV_1_ expressions for multiple clinical outcomes.

We aimed to compare the predictive abilities of ppFEV_1_, FEV_1_ z-score, FEV_1_.Ht^-2^, FEV_1_.Ht^- 3^, and FEV_1_Q and their respective methods of classification of COPD severity for clinical outcomes in a Norwegian COPD cohort followed for 5 years (short-term) and up to 20.4 years (long-term). The clinical outcomes were all-cause mortality, respiratory mortality, cardiovascular mortality, COPD hospitalization, and pneumonia hospitalization.

## METHODS

### Study population

Trøndelag is a county in central Norway with a homogenous and stable population. The HUNT Study invited the entire adult population (≥20 years) of northern Trøndelag to attend clinical examinations and answer questionnaires. Four surveys have been conducted in the periods of 1984-1986 (HUNT1), 1995-1997 (HUNT2), 2006-2008 (HUNT3), and 2017-2019 (HUNT4)(18).

This study included people aged ≥40 years who participated in HUNT2 (n=44,384, 75.2% participation). A 5% random sample (n=2300) and people reporting asthma related symptoms, diagnosis or use of medication (n=7123) were invited to perform spirometry(19). Participants from rural municipalities and participants having airflow limitation (pre-BD FEV_1_/ forced vital capacity (FVC)<0.75 or ppFEV_1_<80 using the European Coal and Steel Community (ECSC) equations(20)) from urban municipalities were invited to attend post-BD spirometry (n=5678). These airflow limitation criteria were used to allow for future changes in diagnostic and severity classification of COPD. Among those performing post-BD spirometry (n=4178, 73.6% of invited), 3840 (91.9%) had acceptable manoeuvres. There were 1350 people with COPD when we defined COPD as having post-BD FEV_1_/FVC<0.70 (fixed-ratio criteria) and [respiratory symptoms (daily cough in periods, cough with phlegm, wheezing, or dyspnoea) and/or self-reported doctor-diagnosed COPD](1). There were 894 people with COPD when we applied lower limit of normal (LLN) criteria i.e. post-BD FEV_1_/FVC z-scores< -1.645 (13). For the analysis, we included 890 people with COPD who met both the fixed-ratio and LLN criteria so that the same sample was used in all analyses regardless of COPD classification (Supplementary Figure E1).

### Spirometry and lung function classification

Post-BD spirometry was performed 30 minutes after inhalation of 1 mg terbutaline according to the 1994 ATS-guidelines(21, 22). Quality assurance of spirometric measurements is described in detail elsewhere(22, 23).

We defined expressions of FEV_1_ such as ppFEV_1_, FEV_1_ z-score, FEV_1_.Ht^-2^, FEV_1_.Ht^-3^, and FEV_1_Q as suggested by the previous studies(1-4, 6, 9, 11, 12). The Global Lung Function Initiative (GLI)-2012 reference equation was used to calculate ppFEV_1_, ppFVC, FEV_1_ z-scores, and FVC z-scores(13, 22). FEV_1_ was standardized by the square of height in meters to calculate FEV_1_.Ht^-2^ (6, 12) and by the cube of height in meters to calculate FEV_1_.Ht^-3^ (9, 11). FEV_1_ was standardized by sex-specific lowest percentile (0.5L for men and 0.4L for women) of FEV_1_ distribution to calculate FEV_1_Q as suggested by Miller et al. who found this in a large European population consisting of 3 cohorts(9).

We categorized ppFEV_1_ according to the GOLD to define GOLD grades(1) and according to the ATS/ERS to define ATS/ERS grades(2) for the classification of COPD severity. We defined FEV_1_ z-score grades according to the recommendation from Quanjer et. al(4). FEV_1_.Ht^-2^ grades were defined suggested by Miller et. al(6). For FEV_1_.Ht^-3^, and FEV_1_Q, no widely acceptable cut-points for the classification of COPD severity have been recommended(5-7, 9, 10, 14, 15). We generated quartiles of ppFEV_1_, FEV_1_ z-score, FEV_1_.Ht^-2^, FEV_1_.Ht^-3^, and FEV_1_Q distribution from our study cohort for the classification of COPD severity for comparisons.

The different expressions of FEV_1_ and their respective methods of classification of COPD severity are presented in Supplementary Table E1.

### Clinical examination and questionnaires

From clinical examinations and questionnaires, information on age, sex, body mass index (BMI, kg/m^2^), smoking status, smoking packs-years, physical activity, education, diabetes ever, asthma ever, cardiovascular disease, systolic blood pressure (mmHg), and non-fasting total serum cholesterol (mmol/L) were recorded.

Age in years was recorded to one decimal place. Height and weight were measured with light clothing and without shoes. Height was rounded to the nearest centimetre and weight rounded to the half kilogram(19, 24). Cardiovascular disease included self-reported angina pectoris, myocardial infarction, and stroke. From three measurements of systolic blood pressure, the mean of the last two measurements was used(24).

### Follow-up and outcomes

The study outcomes were all-cause mortality, respiratory mortality, cardiovascular mortality, the first unplanned COPD hospitalization, and the first unplanned pneumonia hospitalization. Follow-up for all outcomes began at the date of participation in HUNT2 and ended at the date of the outcome or on right-censoring, whichever was the earlier. Right-censoring events were emigration (n=3) or end of follow-up. For the short-term follow-up, participants were followed for 5 years and for the long-term follow-up the participants were followed until 31 December 2015. There was no other loss to follow-up. Cause-specific mortality and hospitalizations were identified from the international statistical classification of disease and related health problems (ICD) codes in medical records and are presented in the Supplementary Table E2(25). Date of death and hospitalizations was obtained from the Norwegian Cause of Death Registry and The Nord-Trøndelag Hospital Trust, respectively.

### Statistical analysis

Incidence rates of all-cause mortality, respiratory mortality, cardiovascular mortality, COPD hospitalization, and pneumonia hospitalization were calculated. Cumulative incidence curves for all-cause mortality were constructed through Kaplan-Meier estimates and log-rank tests were used to test differences. Cumulative incidence curves were constructed for cause-specific mortality and hospitalization where Fine and Grey(26) methods were used to account for the competing events and Grey tests(26) were used to test differences in cumulative incidence curves. For respiratory mortality, deaths from other causes were considered competing events. Likewise, for cardiovascular mortality, deaths from other causes were considered competing events. For hospitalization (COPD hospitalization or pneumonia hospitalization), all-cause mortality was considered a competing event. The classifications of COPD severity were used as continuous variables to test for trends with smoking status, the major risk factor for COPD(1).

To assess the association of FEV_1_ expressions with all-cause mortality, we used Cox proportional hazard models to calculate hazard ratios (HRs) and 95% CI. We used Fine and Grey (26) competing risk models for respiratory mortality, cardiovascular mortality, COPD hospitalization, and pneumonia hospitalization. We presented crude HRs and adjusted HRs. The models were adjusted for age (as a continuous variable), sex (women, men), smoking [never, former (<10, 10-19, ≥20 pack-years), current (<10, 10-19, ≥20 pack-years), unknown], body mass index (<25.0, 25.0-29.9, ≥30.0, unknown), and education (<10, ≥10 years, unknown).

Proportional hazard assumptions were tested through log-log survival curves and Schoenfeld residuals tests(27). Multicollinearity was tested where the variance inflation factor (VIF) was less than 1.2 in all models(28, 29).

A regression tree method(30) that accounts for time and multiple outcomes was applied to obtain optimal cut-offs of FEV_1_ expressions. We defined models for each FEV_1_ expression in predicting multiple outcomes such as respiratory mortality, cardiovascular mortality, other cause-related mortality, and COPD hospitalization over the follow-up time.

The clinical outcomes are time-dependent, for example a healthy person may have disease over the follow-up time. Hence, we applied incident/dynamic time-dependent area under the receiver operating characteristic curves (AUCs) that accounts for time in order to compare the predictive abilities of FEV_1_ expressions and their respective methods of classification of COPD severity to predict all-cause mortality, respiratory mortality, cardiovascular mortality, COPD hospitalization, and pneumonia hospitalization(31-34). For cause-specific mortality and hospitalization, AUCs accounting for competing risks were calculated(33). We used crude models to compare AUCs because the clinical decision does not explicitly take into account other factors(9). We used 10,000 bootstrap iterations to calculate 95% CI for AUCs(35). A general bootstrap algorithm (gBA)(36) was applied to compare the AUCs.

Statistical analysis was performed using R 3.6.1 software (http://www.r-project.org).

### Replication cohort

We used the UK Biobank as a replication cohort. Here, 6495 people with COPD were followed for 5 years to investigate the predictive abilities of FEV_1_ expressions and their respective methods of classification of COPD severity to predict all-cause, respiratory, and cardiovascular mortality. The optimal cut-offs of FEV_1_Q generated in HUNT were tested in UK Biobank. The details of the COPD cohort from UK Biobank are presented in the Supplementary Text E1.

### Ethics

Ethical approval was obtained from the Regional Committees for Medical and Health Research Ethics (2015/1461/REK midt). All participants gave informed written consent.

## RESULTS

This population-based COPD cohort (n=890) was followed-up for 5 years (short-term) and up to 20.4 years (long-term). During the long-term follow-up period, 615, 195 and 184 died due to all-causes, respiratory, and cardiovascular diseases, respectively, and 428 and 311 were hospitalized due to COPD and pneumonia, respectively. At baseline, the average age of participants was 63.8 years, six out of ten participants were men, and more than half were current smokers (Table 1, Supplementary Table E3). A trend for increasing mean of smoking pack-years (except for FEV_1_.Ht^- 2^ grades, p value=0.062) and increasing cumulative incidence of all-cause mortality, respiratory mortality, cardiovascular mortality (only for FEV_1_.Ht^-2^ quartiles, FEV_1_.Ht^-3^ quartiles, and FEV_1_Q quartiles), COPD hospitalization, and pneumonia hospitalization was observed with worsening categories of classifications of COPD severity (Table 1, Figure 1). Similar results were observed in cumulative incidence curves (Figure 2, Supplementary Figure E2-5).

**Table 1.**
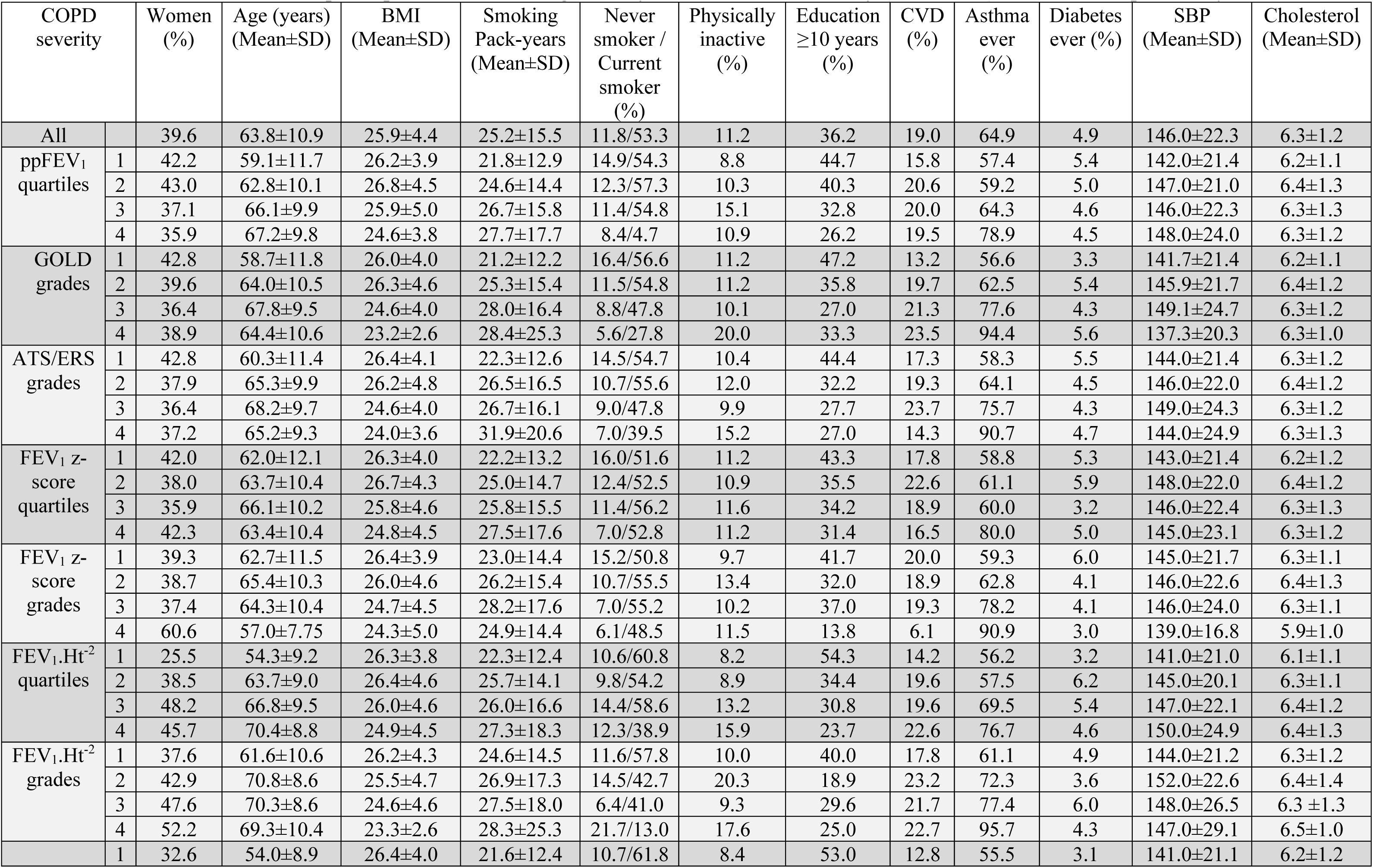
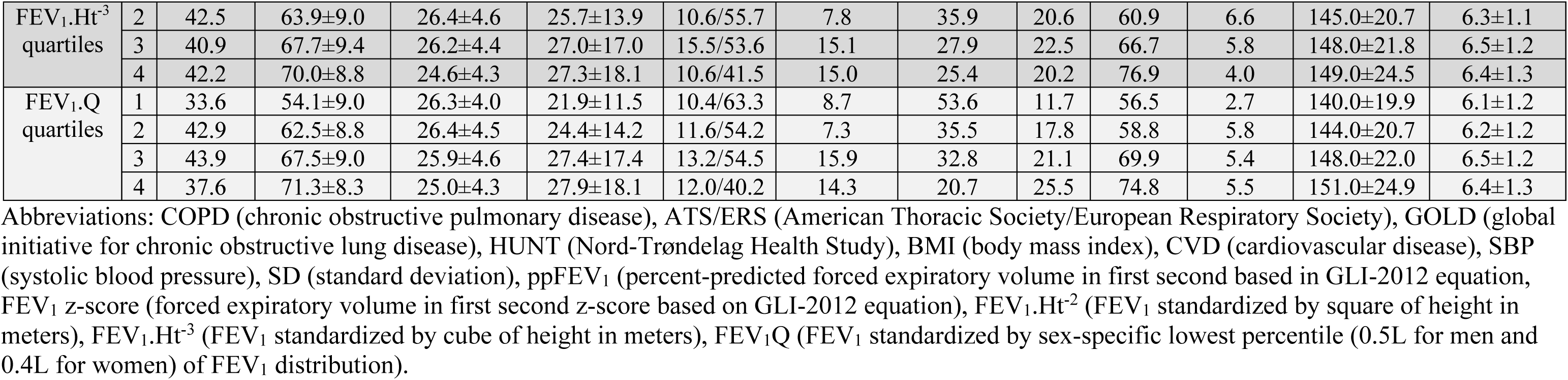
Baseline characteristics of participants with COPD aged ≥40 years in the HUNT2 study (1995-1997) followed for up to 20.4 years.

**Figure 1.**
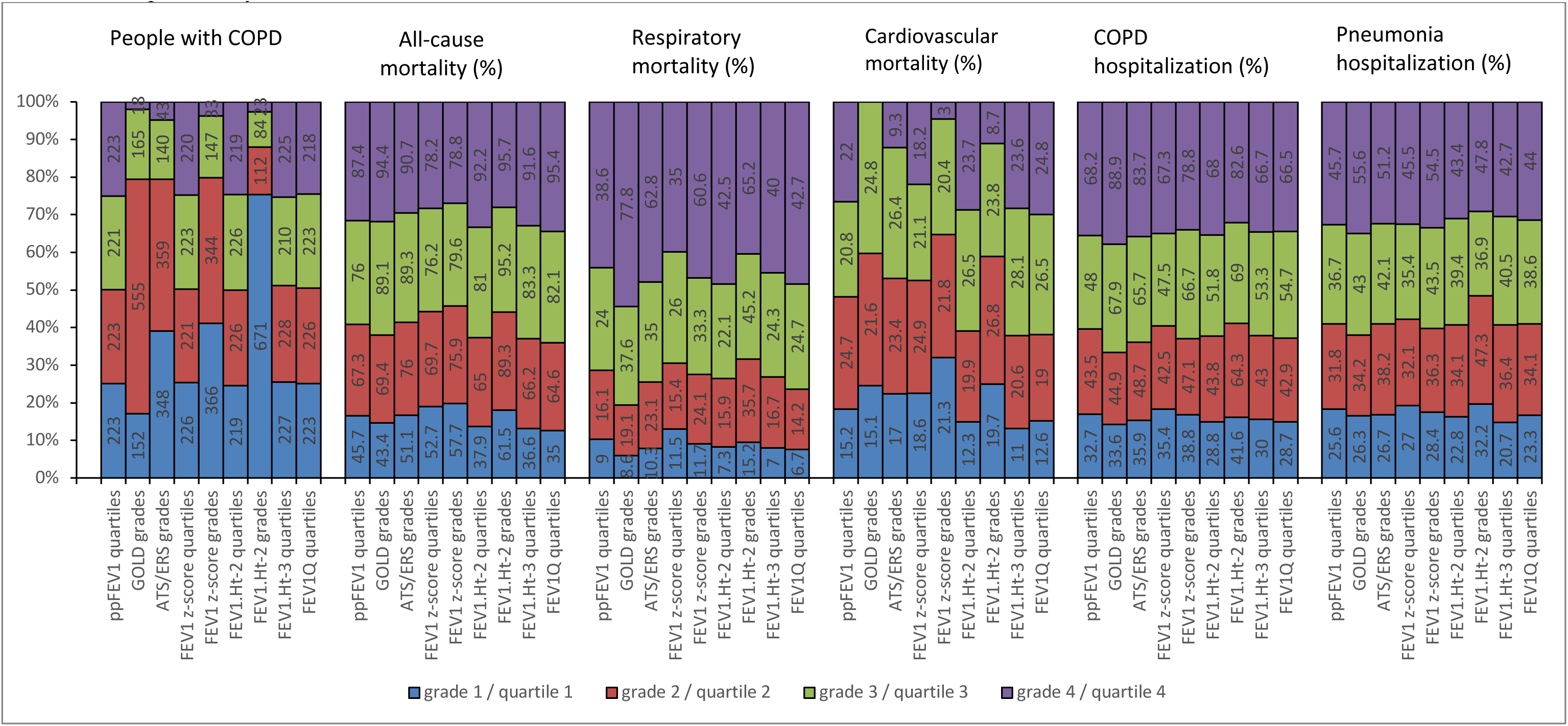
Distribution of participants with COPD, all-cause mortality, respiratory mortality, cardiovascular mortality, COPD hospitalization, and pneumonia hospitalization in categories of ppFEV_1_ quartiles, GOLD grades, ATS/ERS grades, FEV_1_ z-score quartiles, FEV_1_ z-score grades, FEV_1_.Ht^-2^ quartiles, FEV_1_.Ht^-2^ grades, FEV_1_.Ht^-3^ quartiles, and FEV_1_Q quartiles among participants with COPD aged ≥40 years in the HUNT2 study (1995-1997) followed for up to 20.4 years. Abbreviations: COPD (chronic obstructive pulmonary disease), ATS/ERS (American Thoracic Society/European Respiratory Society), GOLD (global initiative for chronic obstructive lung disease), HUNT (Nord-Trøndelag Health Study), ppFEV_1_ (percent-predicted forced expiratory volume in first second based in GLI-2012 equation, FEV_1_ z-score (forced expiratory volume in first second z-score based on GLI-2012 equation), FEV_1_.Ht^-2^ (FEV_1_ standardized by square of height in meters), FEV_1_.Ht^-3^ (FEV_1_ standardized by cube of height in meters), FEV_1_Q (FEV_1_ standardized by sex-specific lowest percentile (0.5L for men and 0.4L for women) of FEV_1_ distribution).

**Figure 2.**
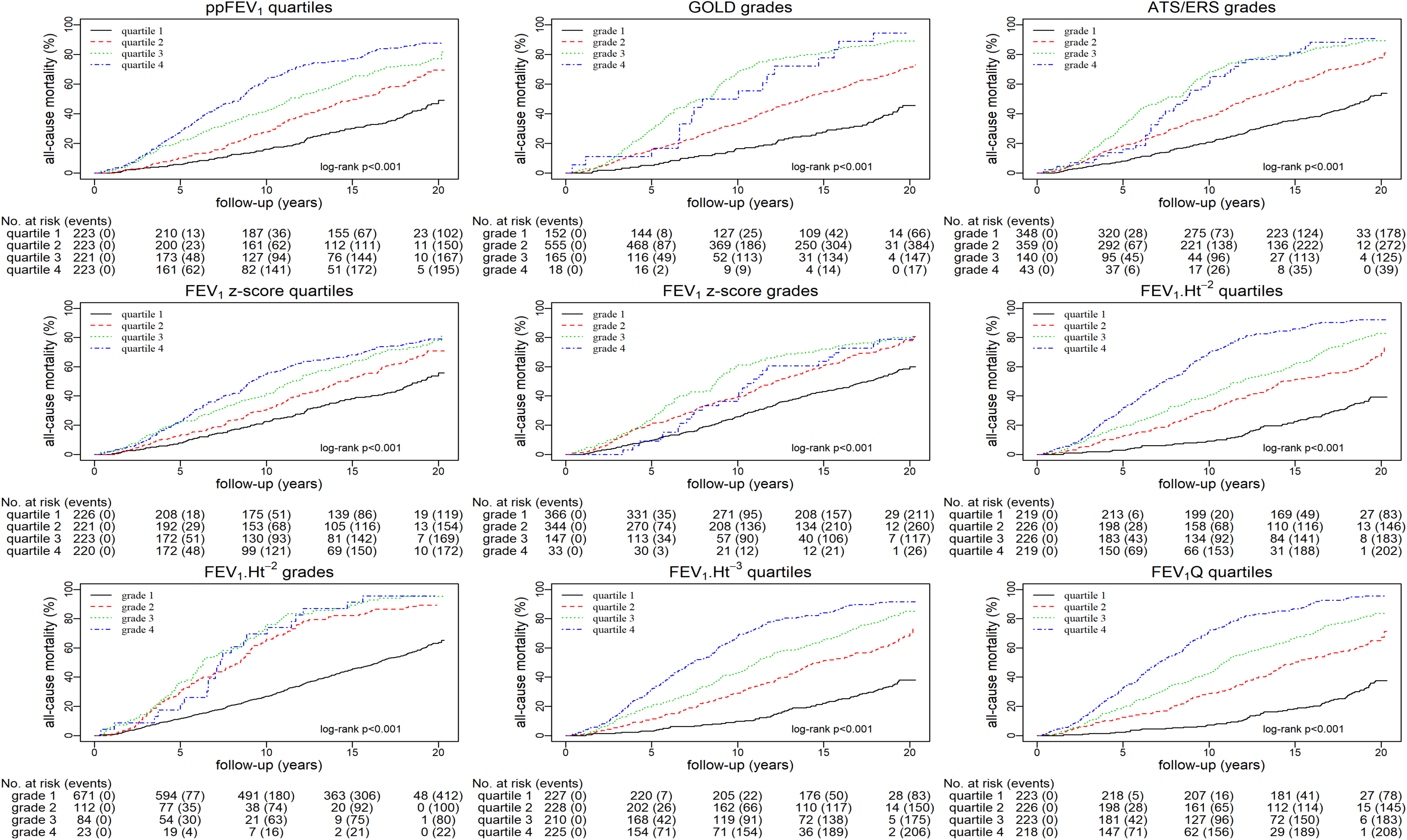
Cumulative incidence curves of classifications of COPD severity for all-cause mortality among participants with COPD aged ≥40 years in the HUNT2 study (1995-1997) followed for up to 20.4 years.

In long-term follow-up, the HRs (95% CI) for all-cause mortality for the lowest compared to the highest grade or quartile were 3.97 (3.11-5.05) for ppFEV_1_ quartiles, 4.29 (2.51-7.32) for GOLD grades, 3.43 (2.42-4.87) for ATS/ERS grades, 2.32 (1.83-2.93) for FEV_1_ z-score quartiles, 1.77 (1.18-2.66) for FEV_1_ z-score quartiles, 6.68 (5.15-8.68) for FEV_1_.Ht^-2^ quartiles, 3.44 (2.23-5.30) for FEV_1_.Ht^-2^ grades, 6.71 (5.17-8.70) for FEV_1_.Ht^-3^ quartiles, and 8.48 (6.49-11.08) for FEV_1_Q (Figure 3). Similarly, for cause-specific mortality and hospitalization, the HRs of the lowest quartile of FEV_1_Q was higher than those of the lowest quartile of ppFEV_1_ (Figure 3). Generally, similar results were observed in adjusted models (Supplementary Figure E6).

**Figure 3.**
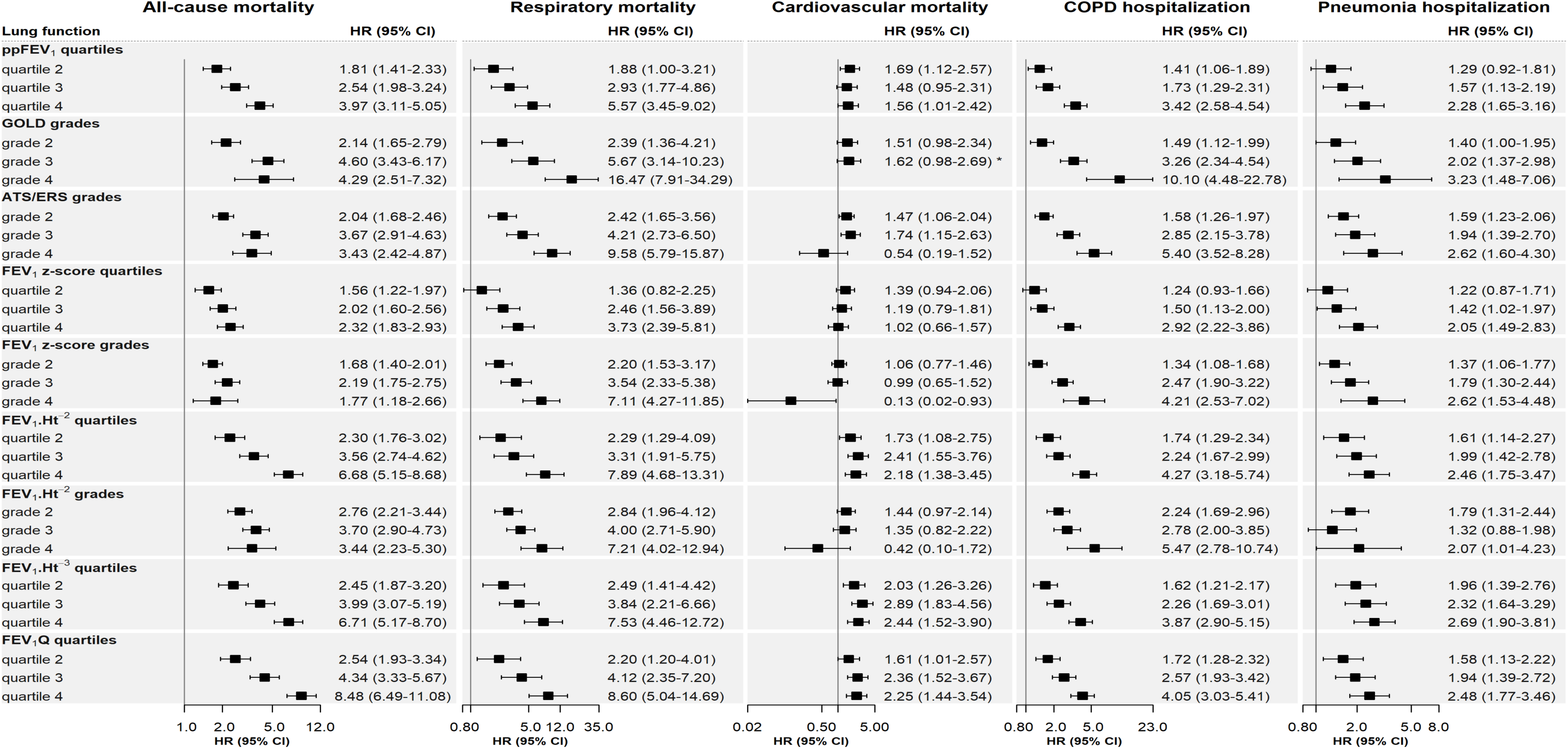
Crude hazard ratios for different expressions of FEV_1_ and their respective methods of classification of COPD severity for all-cause mortality, respiratory mortality, cardiovascular mortality, COPD hospitalization, and pneumonia hospitalization among participants with COPD aged ≥40 years in the HUNT2 study (1995-1997) followed for up to 20.4 years. Abbreviations: COPD (chronic obstructive pulmonary disease), ATS/ERS (American Thoracic Society/European Respiratory Society), GOLD (global initiative for chronic obstructive lung disease), HUNT (Nord-Trøndelag Health Study), CI (confidence interval), HR (Hazard ratio), ppFEV_1_ (percent-predicted forced expiratory volume in first second based in GLI-2012 equation, FEV_1_ z-score (forced expiratory volume in first second z-score based on GLI-2012 equation), FEV_1_.Ht^-2^ (FEV_1_ standardized by square of height in meters), FEV_1_.Ht^-3^ (FEV_1_ standardized by cube of height in meters), FEV_1_Q (FEV_1_ standardized by sex-specific lowest percentile (0.5L for men and 0.4L for women) of FEV_1_ distribution). *-grades/quartiles 3-4 were combined due to zero cases in grade/quartile 4

When using FEV_1_expressions as continuous measures, the short-term AUCs (95% CI) for all-cause mortality were 64.5 (60.6-68.3) for ppFEV_1_, 58.8 (55.1-62.4) for FEV_1_z-score, 68.9 (65.4-72.4) for FEV_1_.Ht^-2^, 69.3 (65.7-72.7) for FEV_1_.Ht^-3^, and 70.2 (66.6-73.6) for FEV_1_Q (Figure 4). The corresponding long-term AUCs (95% CI) were 61.9 (58.9-64.6), 57.9 (55.4-60.5), 66.8 (62.6-70.4), 66.6 (62.5-70.1), and 68.3 (64.0-72.1), respectively (Figure 5). Similarly, the short-term and long-term AUCs for cause-specific mortality and hospitalization for FEV_1_Q were the highest compared to ppFEV_1_, FEV_1_ z-score, FEV_1_.Ht^-2^, and FEV_1_.Ht^-3^ (Figure 4, Figure 5).

**Figure 4.**
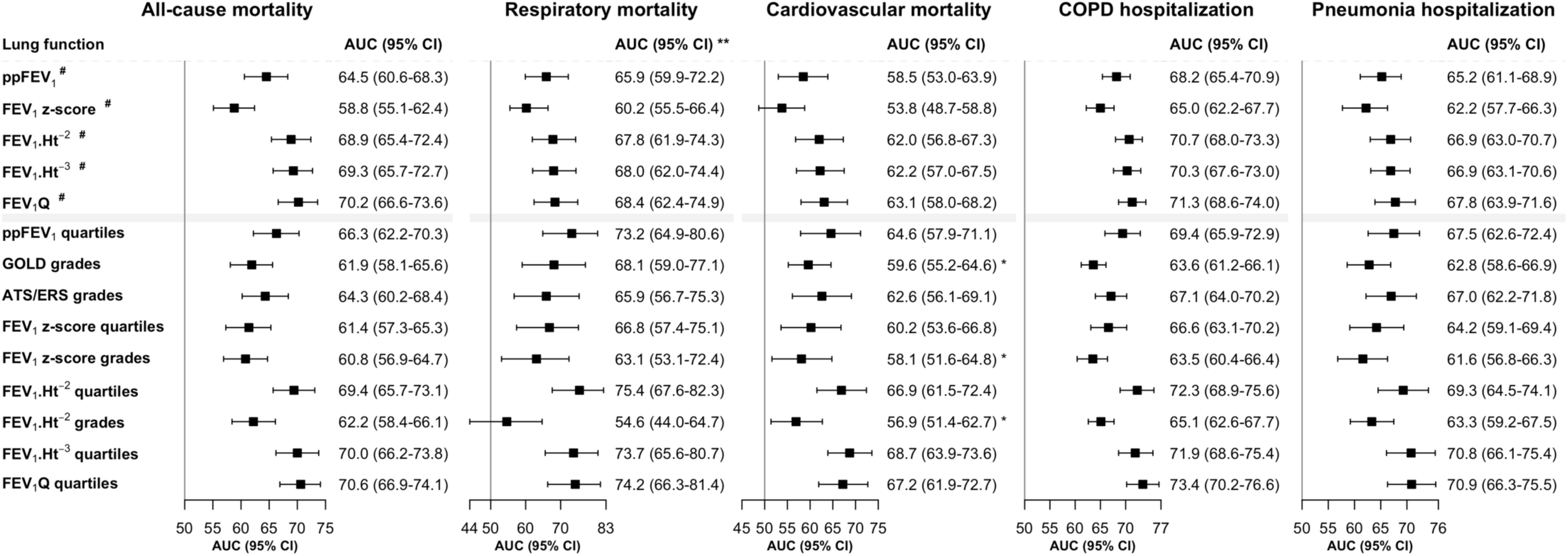
The AUCs for different expressions of FEV_1_ and their respective methods of classification of COPD severity for all-cause mortality, respiratory mortality, cardiovascular mortality, COPD hospitalization, and pneumonia hospitalization among participants with COPD aged ≥40 years in the HUNT2 study (1995-1997) followed for 5 years. Abbreviations: COPD (chronic obstructive pulmonary disease), ATS/ERS (American Thoracic Society/European Respiratory Society), GOLD (global initiative for chronic obstructive lung disease), HUNT (Nord-Trøndelag Health Study), CI (confidence interval), AUC (area under receiver operating characteristics curves), ppFEV_1_ (percent-predicted forced expiratory volume in first second based in GLI-2012 equation, FEV_1_ z-score (forced expiratory volume in first second z-score based on GLI-2012 equation), FEV_1_.Ht^-2^ (FEV_1_ standardized by square of height in meters), FEV_1_.Ht^-3^ (FEV_1_ standardized by cube of height in meters), FEV_1_Q (FEV_1_ standardized by sex-specific lowest percentile (0.5L for men and 0.4L for women) of FEV_1_ distribution). _#_-continuous variables, *-grades/quartiles 3-4 were combined due to zero cases in grade/quartile 4, **-grades/quartiles 2-4 were analysed due to zero cases in grade/quartile 1 of GOLD grades, FEV_1_.Ht^-2^ quartiles, FEV_1_.Ht^-3^ quartiles, FEV_1_Q quartiles. **-Note: similar differences in AUCs were observed when grade/quartile 1-2 were combined for respiratory mortality.

**Figure 5.**
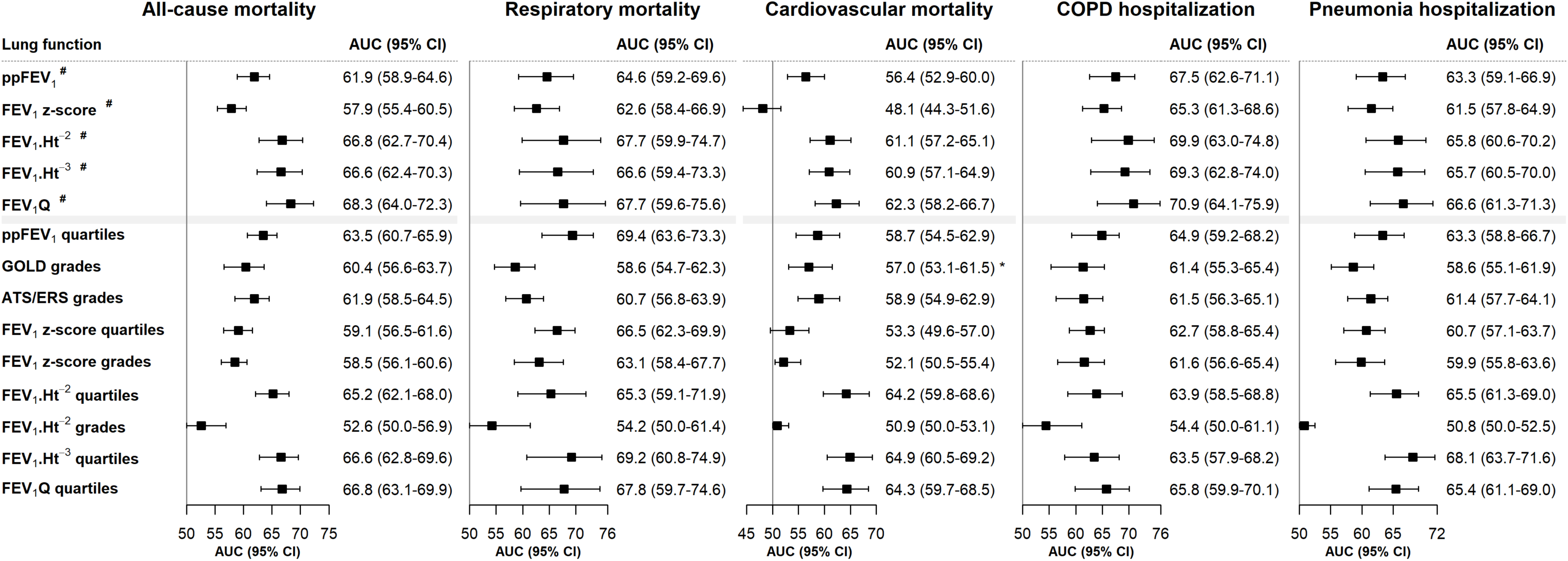
The AUCs for different expressions of FEV_1_ and their respective methods of classification of COPD severity for all-cause mortality, respiratory mortality, cardiovascular mortality, COPD hospitalization, and pneumonia hospitalization among participants with COPD aged ≥40 years in the HUNT2 study (1995-1997) followed for up to 20.4 years. Abbreviations: COPD (chronic obstructive pulmonary disease), ATS/ERS (American Thoracic Society/European Respiratory Society), GOLD (global initiative for chronic obstructive lung disease), HUNT (Nord-Trøndelag Health Study), CI (confidence interval), AUC (area under receiver operating characteristics curves), ppFEV_1_ (percent-predicted forced expiratory volume in first second based in GLI-2012 equation, FEV_1_ z-score (forced expiratory volume in first second z-score based on GLI-2012 equation), FEV_1_.Ht^-2^ (FEV_1_ standardized by square of height in meters), FEV_1_.Ht^-3^ (FEV_1_ standardized by cube of height in meters), FEV_1_Q (FEV_1_ standardized by sex-specific lowest percentile (0.5L for men and 0.4L for women) of FEV_1_ distribution). *-grades/quartiles 3-4 were combined due to zero cases in grade/quartile 4.

When using FEV_1_ expressions as classifications of COPD severity, the short-term and long-term AUCs for COPD classifications based on reference-independent lung functions (FEV_1_.Ht^-2^ quartiles, FEV_1_.Ht^-3^ quartiles, and FEV_1_Q quartiles) were higher than COPD classifications based on reference-dependent lung functions (ppFEV_1_ quartiles, GOLD grades, ATS/ERS grades, FEV_1_ z-score quartiles, and FEV_1_ z-score grades) in predicting all-cause mortality, respiratory mortality, cardiovascular mortality, COPD hospitalization, and pneumonia hospitalization (Figure 4, Figure 5). However, the AUC for FEV_1_.Ht^-2^ grades based on reference-independent lung function (FEV_1_.Ht^-2^) was generally less than GOLD grades that is based on reference-dependent lung function (ppFEV_1_) (Figure 4, Figure 5).

Additionally, we identified optimal cut-offs of FEV_1_ expressions that are reference-independent through a regression tree approach (Supplementary Figure E7). The short-term and long-term AUCs for optimal cut-offs of FEV_1_Q (2.8, 4.1, 5.2) termed as FEV_1_Q grades (Figure 6) were the highest compared to optimal cut-offs of FEV_1_.Ht^-2^ and FEV_1_.Ht^-3^ to predict clinical outcomes studied (data not shown). The FEV_1_Q grades captured more very severe cases compared to GOLD grades (Figure 1, Figure 6). Generally, in long-term follow-up the HRs for the lowest grade of FEV_1_Q were higher than those of the lowest grade of GOLD in predicting clinical outcomes studied (Figure 3, Figure 6). The short-term and long-term AUCs of FEV_1_Q grades were higher than those of the GOLD grades (p value <0.001) (Figure 4, Figure 5, Figure 6).

**Figure 6.**
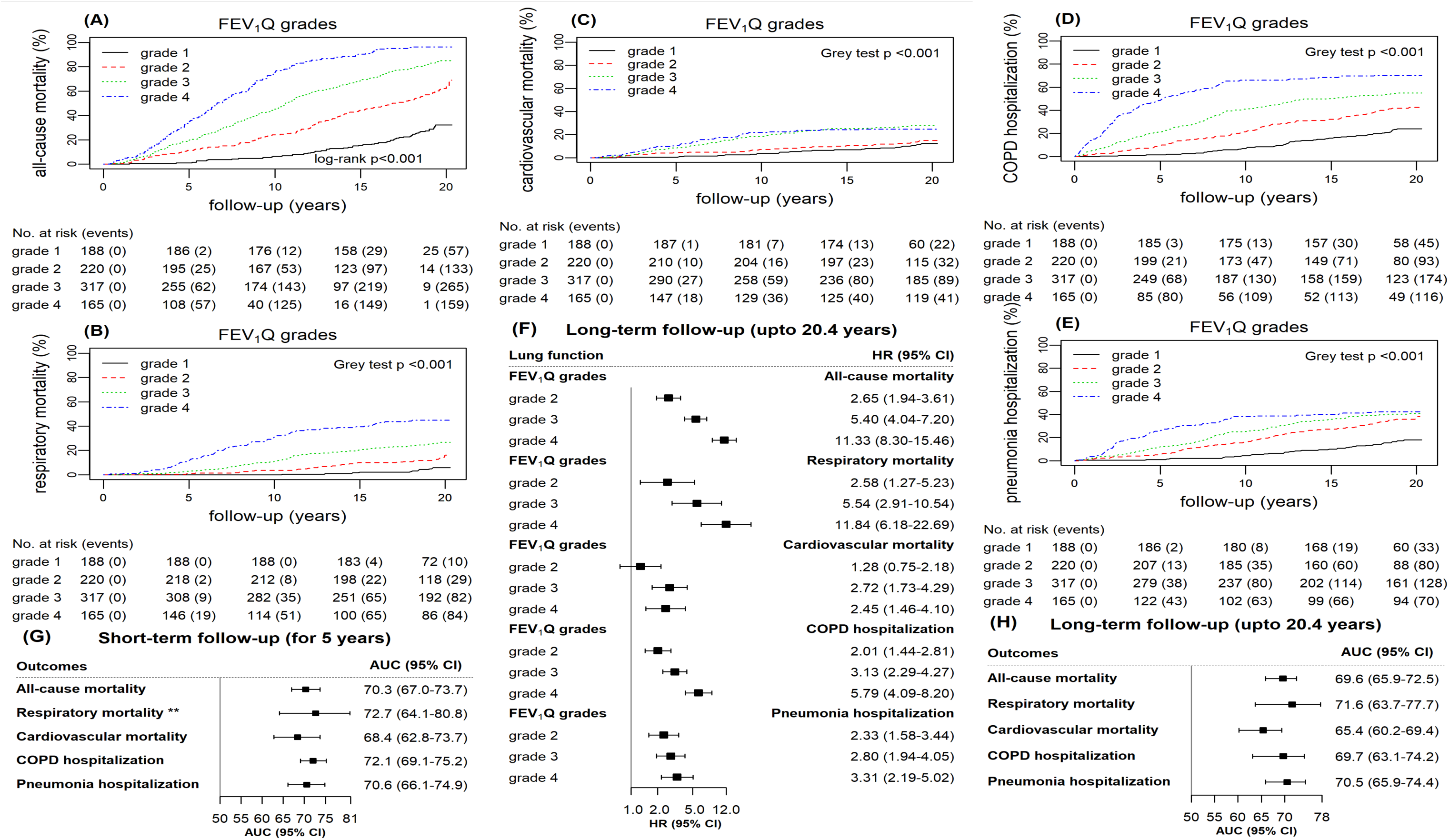
The cumulative incidence curves (A-E), hazard ratios (F), and AUCs (G-H) for optimal cut-offs of FEV_1_Q (FEV_1_Q grades) for all-cause mortality, respiratory mortality, cardiovascular mortality, COPD hospitalization, and pneumonia hospitalization among participants with COPD aged ≥40 years in the HUNT2 study (1995-1997) followed for 5 years and up to 20.4 years. Abbreviations: COPD (chronic obstructive pulmonary disease), ATS/ERS (American Thoracic Society/European Respiratory Society), GOLD (global initiative for chronic obstructive lung disease), HUNT (Nord-Trøndelag Health Study), CI (confidence interval), HR (Hazard ratio), AUC (area under receiver operating characteristics curves), FEV_1_ (forced expiratory volume in first second), FEV_1_Q (FEV_1_ standardized by sex-specific lowest percentile (0.5L for men and 0.4L for women) of FEV_1_ distribution). **-FEV_1_Q grades 2-4 were analysed due to zero cases in grade 1. **-Note: similar differences in AUCs were observed when grade 1-2 were combined for respiratory mortality.

### Replication cohort

Similar results were found in a COPD cohort from UK Biobank followed for 5 years (Supplementary Figure E8 and Figure E9). The short-term AUC for FEV_1_Q as a continuous variable or FEV_1_Q grades as classification of COPD severity was the highest for all-cause mortality, respiratory mortality, and cardiovascular mortality (Supplementary Figure E9).

## DISCUSSION

In this population-based study, we found that among all FEV_1_ expressions, FEV_1_Q was the best predictor of clinical outcomes such as all-cause mortality, respiratory mortality, cardiovascular mortality, COPD hospitalization, and pneumonia hospitalization, followed by FEV_1_.Ht^-2^ or FEV_1_.Ht^-3^ across 5 years and up to 20.4 years of follow-up. The optimal cut-offs of FEV_1_Q (FEV_1_Q grades) have substantially higher short-term and long-term predictive abilities compared to GOLD grades in predicting clinical outcomes.

Others have observed similar results for all-cause mortality(5-7, 9, 10, 14, 15), exacerbation(5), and all-cause hospitalization(14, 15). We observed that compared to the highest quartile, the risk of all-cause mortality for the lowest quartiles of FEV_1_Q (adjusted HR 3.18) was higher than for the lowest quartile of ppFEV_1_ (adjusted HR 2.65) and other FEV_1_ expressions. Pedone et al.(7) observed similar results, where adjusted HR for all-cause mortality of lowest quintile was 4.45 for FEV_1_Q and 3.28 for ppFEV_1_ compared to the highest quintile. Miller et al.(9) observed a similar result where the risk of all-cause mortality for the lowest decile versus the highest decile of FEV_1_Q (adjusted HR 6.9) was higher than for the ppFEV_1_ (adjusted HR 4.1). Until now, no studies have investigated cause-specific mortality and hospitalization in this context. We observed an adjusted HR of 3.42 for FEV_1_Q and 2.66 for ppFEV_1_ for the risk of COPD hospitalization in the lowest compared to the highest quartile. Huang et al.(5) reported similar results for exacerbation. Although COPD hospitalization and exacerbation are related, it is important to note that most exacerbations do not lead to hospitalizations.

In our study, we found that FEV_1_Q, which is independent of reference values, was the best predictor for clinical outcomes studied. Participants with COPD in this population-based cohort (HUNT Study) and replication cohort (UK Biobank) had mean age of 63.8 and 57.5, respectively, and included 18 (2.0%) and 108 (1.7%) very severe cases (GOLD grades 4), respectively, which might best represent a primary health care setting. Our study findings corresponds to the finding of Miller et al.(9) and others(5, 7, 15) including either population-based or hospital-based cohorts. Additionally, only Miller et al.(9) and Hegendorfer et al.(15) have investigated the predictive ability of FEV_1_ expressions as continuous measures. Miller et al.(9) studied both population-based and hospital-based cohorts and observed that the ability to predict mortality was the highest for FEV_1_Q (AUC 63.1) compared to ppFEV_1_ (AUC 58.6) and other FEV_1_ expressions, where they used ECSC reference equation(20) for the calculation of ppFEV_1_. Our finding is also supported by Hegendorfer et al.(15) who studied a population-based cohort and used the Concordance index and risk classification improvement measures and observed that FEV_1_.Ht^-3^ and FEV_1_Q were better in predicting mortality, all-cause hospitalization, mental and physical decline. However, they studied people aged ≥80 years with only 14% of participants having asthma and/or COPD. Other studies have investigated the prediction performance of categorical FEV_1_ expressions(5, 7). Huang et al.(5) studied a hospital-based cohort and found FEV_1_Q as quartiles was the best predictor of mortality and exacerbation in a limited sample of 296 people with COPD. Pedone et al.(7) studied hospital-based cohort and used concordance index and observed similar results to ours for mortality when FEV_1_ expressions were categorized into quintiles among a limited sample of 318 people with COPD aged ≥65 years.

FEV_1_ is a continuous variable where, the expression of FEV_1_ is used for indicating lung function impairments in respiratory medicine and ppFEV_1_ is most commonly used for this purpose(1). Furthermore, the classification of COPD severity based on FEV_1_ expression has been clinically used for guiding therapy and predicting the outcomes of COPD patients(1). The GOLD grades based on ppFEV_1_, have been widely used for clinical purposes in classifying COPD severity(1). However, they have been criticized due to their susceptibility to physiological variation and poor prediction ability(3-6). The FEV_1_ z-score avoids this bias due to physiological variation(3, 4). Vaz Fragoso et al.(3) used the reference equation from NHANES III(37) and found that severe COPD based on FEV_1_ z-score was associated with high risk of respiratory symptoms and death. Tejero et al.(16) found that FEV_1_ z-score predicted mortality worse than ppFEV_1_ where they calculated the predicted values from GLI reference equation(13). The ppFEV_1_ and FEV_1_ z-score are based on reference values and depend on the choice of reference equation(13, 22, 38, 39). Furthermore, the performance of the methods of classification of COPD severity based on these FEV_1_ expressions might vary with reference values. Miller et al.(6, 9, 10) found that the FEV_1_ expressions such as FEV_1_.Ht^-2^, FEV_1_.Ht^-3^, and FEV_1_Q, which are independent of reference equations, were better correlated with mortality than those that are dependent on reference equations. Additionally, Miller et al.(9) found that FEV_1_Q predicted mortality better than other FEV_1_ expressions. Extending this knowledge, our study investigated the short-term and long-term predictive ability for several clinical outcomes supporting FEV_1_Q as a stronger predictor than other FEV_1_ expressions. This indicates that the severity in people with COPD appears to be better related to how far the FEV_1_ of that person is from “bottom line” rather than how far it is from a “predicted value”.

The predictive ability of a classification of COPD severity based on a FEV_1_ expression largely depends on the choice of cut-offs. For example, the GOLD grades, ATS/ERS grades, and ppFEV_1_ quartiles had different predictive abilities in our study even though all are derived from ppFEV_1_. Huang et al(5) observed similar results. Therefore, the optimal cut-offs of FEV_1_ expressions for classification of COPD severity were investigated in this study and we found that cut-offs for FEV_1_Q (2.8, 4.1, 5.2, FEV_1_Q grades) were generally best in predicting short-term and long-term clinical outcomes. The optimal cut-offs should be further investigated in a large multi-ethnic population with a wide age range. In a clinical setting, information such as age, sex, and height of COPD patients is easily available. Therefore, using FEV_1_Q (or other expressions of FEV_1_ that are independent of reference equations) for risk classification of COPD patients might be easy to apply and avoid variation due to dependence on reference equations(9). Furthermore, multidimensional prognostic indices that combine reference independent FEV_1_ expressions with symptoms, exacerbation, risk factors, and/or biomarkers should be investigated further.

This study had several strengths. To our knowledge, this is the first study that has extensively studied the short-term and long-term predictive abilities of a range of FEV_1_ expressions both as categories or quartiles and as continuous measures to predict several clinical outcomes. We had complete information on mortality and there was no loss to follow-up other than very few emigrations (3 out of 890 participants). To reduce measurement error, quality assurance of spirometry curves was performed(22, 23). Notably, we have replicated our results in a large COPD cohort from UK Biobank.

This study also had certain limitations. We had information on COPD hospitalizations only from the hospitals in the study area (northern Trøndelag) and lacked data from other hospitals in Norway. Additionally, there was missing information on some covariates, therefore, to avoid sample loss in adjusted models, a missing indicator variable (missing information as ‘unknown’ category) was used which might bias the associations between the FEV_1_ expressions and the clinical outcomes studied. Our methods may not capture non-linear associations between FEV_1_ expressions and mortality(16) or hospitalization and further studies investigating these approaches are needed.

In summary, FEV_1_Q was the best predictor for clinical outcomes such as all-cause mortality, respiratory mortality, cardiovascular mortality, COPD hospitalization, and pneumonia hospitalization compared to a broad range of commonly applied FEV_1_ expressions. The findings highlight improved prediction of outcomes by use of FEV_1_Q for expressing spirometric lung function impairment and the classification of COPD severity.

## Data Availability

Data are available upon reasonable request and with permission of HUNT Research Centre.

## Author’s contributions

LB, LL, AL and BMB conceived and designed the study. LB analysed the data. LB wrote the first draft of the manuscript. All authors interpreted the results, revised and approved the manuscript for submission. LB and BMB are accountable for the accuracy and integrity of all parts of the work. As project leader for the HUNT2 Lung Study, AL was responsible for planning, data collection and quality assurance of data in the Lung Study.

## Acknowledgements

The Nord-Trøndelag Health Study (HUNT) is a collaboration between HUNT Research Centre (Faculty of Medicine and Health Science, Norwegian University of Science and Technology NTNU), Nord-Trøndelag County Council and the Norwegian Institute of Public Health. The HUNT2 Lung Study was partly funded through a non-demanding grant from AstraZeneca Norway.

## Reference

1. Global Initiative for Chronic Obstructive Lung Disease. Global strategy for the diagnosis, management, and prevention of chronic obstructive pulmonary diesease, 2019. [Accessed 01 January 2020] Available from URL: http://goldcopd.org/.

2. Pellegrino R, Viegi G, Brusasco V, Crapo RO, Burgos F, Casaburi R, Coates A, van der Grinten CP, Gustafsson P, Hankinson J, Jensen R, Johnson DC, MacIntyre N, McKay R, Miller MR, Navajas D, Pedersen OF, Wanger J. Interpretative strategies for lung function tests. The European respiratory journal 2005; 26: 948–968.

3. Fragoso CA, Concato J, McAvay G, Yaggi HK, Van Ness PH, Gill TM. Staging the severity of chronic obstructive pulmonary disease in older persons based on spirometric Z-scores. Journal of the American Geriatrics Society 2011; 59: 1847–1854.

4. Quanjer PH, Pretto JJ, Brazzale DJ, Boros PW. Grading the severity of airways obstruction: new wine in new bottles. European Respiratory Journal 2014; 43: 505.

5. Huang TH, Hsiue TR, Lin SH, Liao XM, Su PL, Chen CZ. Comparison of different staging methods for COPD in predicting outcomes. The European respiratory journal 2018; 51.

6. Miller MR, Pedersen OF, Dirksen A. A new staging strategy for chronic obstructive pulmonary disease. Int J Chron Obstruct Pulmon Dis 2007; 2: 657–663.

7. Pedone C, Scarlata S, Scichilone N, Forastiere F, Bellia V, Antonelli-Incalzi R. Alternative ways of expressing FEV_1_and mortality in elderly people with and without COPD. European Respiratory Journal 2013; 41: 800.

8. Bhatta L, Leivseth L, Carslake D, Langhammer A, Mai X-M, Chen Y, Henriksen AH, Brumpton BM. Comparison of pre-and post-bronchodilator lung function as predictors of mortality: The HUNT Study. Respirology 2019; 0.

9. Miller MR, Pedersen OF. New concepts for expressing forced expiratory volume in 1 s arising from survival analysis. The European respiratory journal 2010; 35: 873–882.

10. Miller MR, Pedersen OF, Lange P, Vestbo J. Improved survival prediction from lung function data in a large population sample. Respiratory Medicine 2009; 103: 442–448.

11. Fletcher C, Peto R, Tinker C, Speizer FE. The natural history of chronic bronchitis and emphysema. An eight-year study of early chronic obstructive lung disease in working men in London. Oxford University Press, 37 Dover Street, London. W1X 4AH; 1976.

12. Sorlie PD, Kannel WB, O’Connor G. Mortality associated with respiratory function and symptoms in advanced age. The Framingham Study. The American review of respiratory disease 1989; 140: 379–384.

13. Quanjer PH, Stanojevic S, Cole TJ, Baur X, Hall GL, Culver BH, Enright PL, Hankinson JL, Ip MSM, Zheng J, Stocks J, the ERS Global Lung Function Initiative. Multi-ethnic reference values for spirometry for the 3–95 year age range: the global lung function 2012 equations: Report of the Global Lung Function Initiative (GLI), ERS Task Force to establish improved Lung Function Reference Values. The European respiratory journal 2012; 40: 1324–1343.

14. Turkeshi E, Vaes B, Andreeva E, Matheï C, Adriaensen W, Van Pottelbergh G, Degryse J-M. Short-term prognostic value of forced expiratory volume in 1 second divided by height cubed in a prospective cohort of people 80 years and older. BMC Geriatr 2015; 15: 15–15.

15. Hegendorfer E, Vaes B, Andreeva E, Mathei C, Van Pottelbergh G, Degryse JM. Predictive Value of Different Expressions of Forced Expiratory Volume in 1 Second (FEV1) for Adverse Outcomes in a Cohort of Adults Aged 80 and Older. Journal of the American Medical Directors Association 2017; 18: 123–130.

16. Tejero E, Prats E, Casitas R, Galera R, Pardo P, Gavilan A, Martinez-Ceron E, Cubillos-Zapata C, Del Peso L, Garcia-Rio F. Classification of Airflow Limitation Based on z-Score Underestimates Mortality in Patients with Chronic Obstructive Pulmonary Disease. Am J Respir Crit Care Med 2017; 196: 298–305.

17. Vaz Fragoso CA, Concato J, McAvay G, Yaggi HK, Van Ness PH, Gill TM. Staging the Severity of Chronic Obstructive Pulmonary Disease in Older Persons Based on Spirometric z-Scores. Journal of the American Geriatrics Society 2011; 59: 1847–1854.

18. Krokstad S, Langhammer A, Hveem K, Holmen TL, Midthjell K, Stene TR, Bratberg G, Heggland J, Holmen J. Cohort Profile: the HUNT Study, Norway. International journal of epidemiology 2013; 42: 968–977.

19. Bhatta L, Leivseth L, Mai X-M, Chen Y, Henriksen AH, Langhammer A, Brumpton BM. Prevalence and trend of COPD from 1995–1997 to 2006–2008: The HUNT study, Norway. Respiratory Medicine 2018; 138: 50–56.

20. Quanjer PH, Tammeling GJ, Cotes JE, Pedersen OF, Peslin R, Yernault JC. Lung volumes and forced ventilatory flows. European Respiratory Journal 1993; 6: 5.

21. Standardization of Spirometry, 1994 Update. American Thoracic Society. Am J Respir Crit Care Med 1995; 152: 1107-1136.

22. Langhammer A, Johannessen A, Holmen TL, Melbye H, Stanojevic S, Lund MB, Melsom MN, Bakke P, Quanjer PH. Global Lung Function Initiative 2012 reference equations for spirometry in the Norwegian population. The European respiratory journal 2016; 48: 1602–1611.

23. Hankinson JL, Eschenbacher B, Townsend M, Stocks J, Quanjer PH. Use of forced vital capacity and forced expiratory volume in 1 second quality criteria for determining a valid test. The European respiratory journal 2015; 45: 1283–1292.

24. Holmen J, Midthjell K, Krüger Ø, Langhammer A, Holmen TL, Bratberg GH, Vatten L, Lund-Larsen PG. The Nord-Trøndelag Health Study 1995-97 (HUNT 2): Objectives, contents, methods and participation. Norsk Epidemiologi 2003; 13: 19–32.

25. Backman H, Eriksson B, Ronmark E, Hedman L, Stridsman C, Jansson SA, Lindberg A, Lundback B. Decreased prevalence of moderate to severe COPD over 15 years in northern Sweden. Respir Med 2016; 114: 103–110.

26. Fine JP, Gray RJ. A Proportional Hazards Model for the Subdistribution of a Competing Risk. Journal of the American Statistical Association 1999; 94: 496–509.

27. Grambsch PM, Therneau TM. Proportional Hazards Tests and Diagnostics Based on Weighted Residuals. Biometrika 1994; 81: 515–526.

28. Belsley DA, Kuh E, Welsch RE. Regression diagnostics : identifying influential data and sources of collinearity. New York (N.Y.) : Wiley; 1980.

29. O’Brien RM. A Caution Regarding Rules of Thumb for Variance Inflation Factors. Quality & Quantity 2007; 41: 673–690.

30. De’ath G. Multivariate regression trees: a new technique for modeling species–environment relationships. Ecology 2002; 83: 1105–1117.

31. Heagerty PJ, Zheng Y. Survival Model Predictive Accuracy and ROC Curves. Biometrics 2005; 61: 92–105.

32. Kamarudin AN, Cox T, Kolamunnage-Dona R. Time-dependent ROC curve analysis in medical research: current methods and applications. BMC Medical Research Methodology 2017; 17: 53.

33. Saha P, Heagerty PJ. Time-Dependent Predictive Accuracy in the Presence of Competing Risks. Biometrics 2010; 66: 999–1011.

34. Martínez-Camblor P, Pardo-Fernández JC. Smooth time-dependent receiver operating characteristic curve estimators. Stat Methods Med Res 2018; 27: 651–674.

35. Bansal A, Heagerty PJ. A Tutorial on Evaluating the Time-Varying Discrimination Accuracy of Survival Models Used in Dynamic Decision Making. Medical Decision Making 2018; 38: 904–916.

36. Martínez-Camblor P, Corral N. A general bootstrap algorithm for hypothesis testing. Journal of Statistical Planning and Inference 2012; 142: 589–600.

37. Hankinson JL, Odencrantz JR, Fedan KB. Spirometric reference values from a sample of the general U.S. population. American journal of respiratory and critical care medicine 1999; 159: 179–187.

38. Langhammer A, Johnsen R, Gulsvik A, Holmen TL, Bjermer L. Forced spirometry reference values for Norwegian adults: the Bronchial Obstruction in Nord-Trondelag Study. The European respiratory journal 2001; 18: 770–779.

39. Moreira S, Fernandes M, Silva M, Escaleira D, Staats R, Valença J, Barros A, Bárbara C. Comparison of the FEV1 value from five reference equations: ESCS 718393, NHANES and GLI. European Respiratory Journal 2017; 50: P2507.

