## Supplementary for "Spirometric classifications of COPD severity as predictive markers for clinical outcomes: the HUNT Study"

**Figure E1.** Flow chart - Post-bronchodilator spirometry among people aged ≥40 years in the HUNT2 study

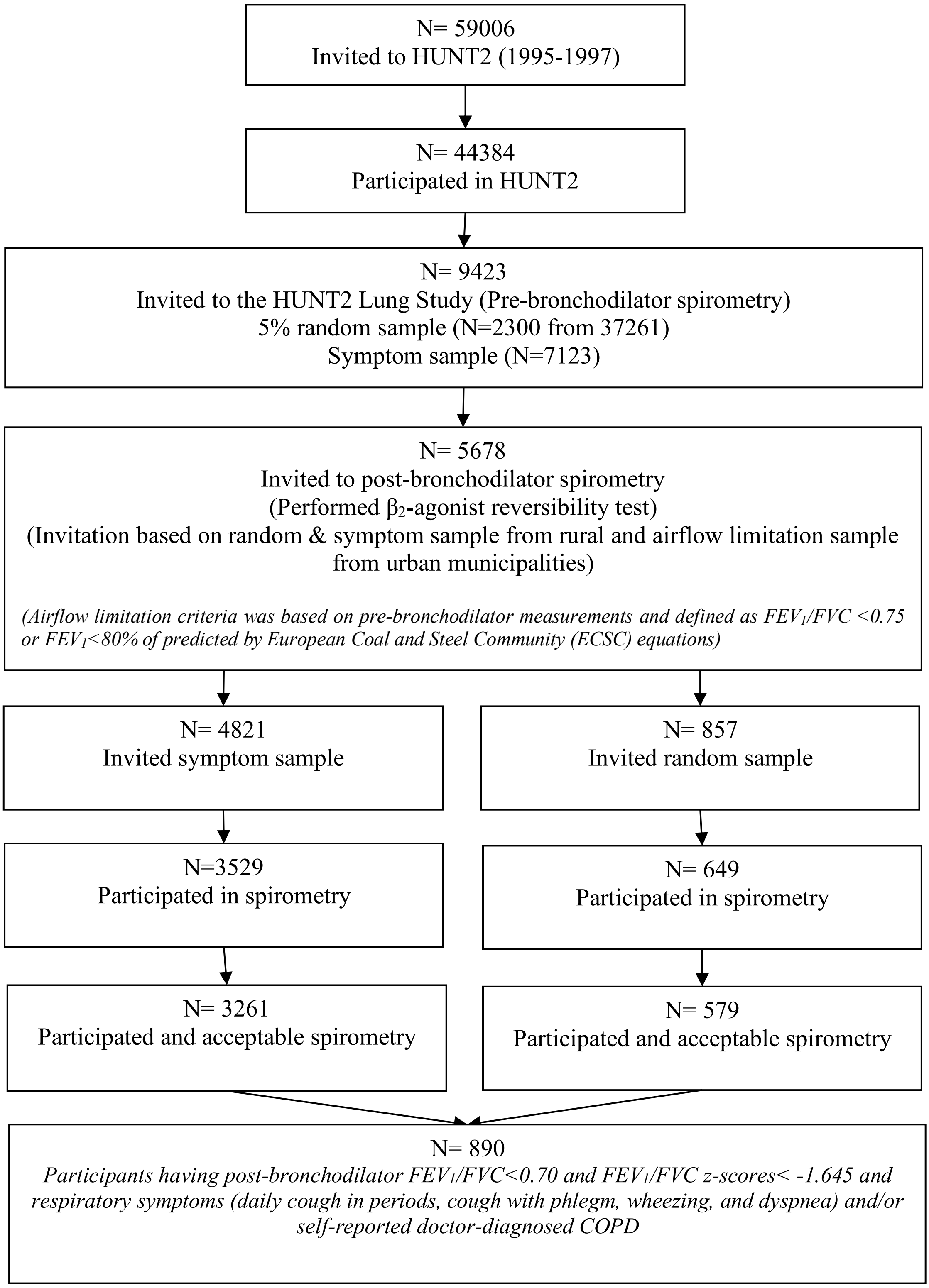

**Table E1.** Different expressions of $FEV_1$ and their respective methods of classification of COPD severity.

| Lung function | | | | | | | | |
|---|---|---|---|---|---|---|---|---|
| $ppFEV_1$ | | | $FEV_1$ z-score | | $FEV_1.Ht^{-2}$ | | $FEV_1.Ht^{-3}$ | $FEV_1Q$ |
| Classification of COPD severity | | | | | | | | |
| $ppFEV_1$ quartiles | GOLD grades | ATS/ERS grades | $FEV_1$ z-score quartiles | $FEV_1$ z-score grades | $FEV_1.Ht^{-2}$ quartiles | $FEV_1.Ht^{-2}$ grades | $FEV_1.Ht^{-3}$ quartiles | $FEV_1Q$ quartiles |
| **quartile 1** ($ppFEV_1 \geq 75.67$) | **grade 1** ($ppFEV_1 \geq 80$) | **grade 1** ($ppFEV_1 \geq 70$) | **quartile 1** ($FEV_1$ z-score $\geq -1.59$) | **grade 1** ($FEV_1$ z-score $\geq -2.0$) | **quartile 1** ($FEV_1.Ht^{-2} \geq 0.79$) | **quartile 1** ($FEV_1.Ht^{-2} \geq 0.5$) | **quartile 1** ($FEV_1.Ht^{-3} \geq 0.46$) | **quartile 1** ($FEV_1Q \geq 4.97$) |
| **quartile 2** ($75.67 > ppFEV_1 \geq 65.40$) | **grade 2** ($80 > ppFEV_1 \geq 50$) | **grade 2** ($70 > ppFEV_1 \geq 50$) | **quartile 2** ($-1.59 > FEV_1$ z-score $\geq -2.19$) | **grade 2** ($-2.0 > FEV_1$ z-score $\geq -3.0$) | **quartile 2** ($0.79 > FEV_1.Ht^{-2} \geq 0.64$) | **quartile 2** ($0.5 > FEV_1.Ht^{-2} \geq 0.4$) | **quartile 2** ($0.46 > FEV_1.Ht^{-3} \geq 0.38$) | **quartile 2** ($4.97 > FEV_1Q \geq 3.96$) |
| **quartile 3** ($65.40 > ppFEV_1 \geq 52.82$) | **grade 3** ($50 > ppFEV_1 \geq 30$) | **grade 3** ($50 > ppFEV_1 \geq 35$) | **quartile 3** ($-2.19 > FEV_1$ z-score $\geq -2.84$) | **grade 3** ($-3.0 > FEV_1$ z-score $\geq -4.0$) | **quartile 3** ($0.64 > FEV_1.Ht^{-2} \geq 0.50$) | **quartile 3** ($0.4 > FEV_1.Ht^{-2} \geq 0.3$) | **quartile 3** ($0.38 > FEV_1.Ht^{-3} \geq 0.30$) | **quartile 3** ($3.96 > FEV_1Q \geq 3.07$) |
| **quartile 4** ($ppFEV_1 < 52.82$) | **grade 4** ($ppFEV_1 < 30$) | **grade 4** ($ppFEV_1 < 35$) | **quartile 4** ($FEV_1$ z-score $< -2.84$) | **grade 4** ($FEV_1$ z-score $< -4.0$) | **quartile 4** ($FEV_1.Ht^{-2} < 0.50$) | **quartile 4** ($FEV_1.Ht^{-2} < 0.3$) | **quartile 4** ($FEV_1.Ht^{-3} < 0.30$) | **quartile 4** ($FEV_1Q < 3.07$) |

Abbreviations: COPD (chronic obstructive pulmonary disease), ATS/ERS (American Thoracic Society/European Respiratory Society), GOLD (global initiative for chronic obstructive lung disease), $ppFEV_1$ (percent-predicted forced expiratory volume in first second based in GLI-2012 equation, $FEV_1$ z-score (forced expiratory volume in first second z-score based on GLI-2012 equation), $FEV_1.Ht^{-2}$ ($FEV_1$ standardized by square of height in meters), $FEV_1.Ht^{-3}$ ($FEV_1$ standardized by cube of height in meters), $FEV_1Q$ ($FEV_1$ standardized by sex-specific lowest percentile (0.5L for men and 0.4L for women) of $FEV_1$ distribution).

**Table E2.** Lists of ICD codes for respiratory mortality, cardiovascular mortality, COPD hospitalization, and pneumonia hospitalization.

| Outcomes | | ICD-10 | ICD-9 |
|---|---|---|---|
| Respiratory mortality | Primary diagnosis | *J00-J99* | *460-519* |
| Cardiovascular mortality | Primary diagnosis | *I00-I99* | *390-459* |
| COPD hospitalization | *- Primary diagnosis | Bronchitis, not specified as acute or chronic * *J40* | Chronic airway obstruction, not elsewhere classified * *496* |
| | | Simple and mucopurulent chronic bronchitis * *J41* | Bronchitis, not specified as acute or chronic * *490* |
| | | Unspecified chronic bronchitis * *J42* | Bronchitis, not specified as acute or chronic * *490* |
| | | Emphysema * *J43* | Emphysema * *492* |
| | | Other chronic obstructive pulmonary disease (COPD) * *J44* | Chronic bronchitis * *491* |
| | #- Primary diagnosis but in combination with a secondary diagnosis of J40-J44 from ICD-10 or 490-492 and 496 from ICD-9 | Influenzas # *J09, J10, J11* | Influenzas # *487, 488* |
| | | Pneumonias # *J12 – J18* | Pneumonias # *480 – 486* |
| | | Dyspnoea # *R06.0* | Shortness of breath # 786.05 |
| | | Acute bronchitis # *J20* | Acute bronchitis # *466.0* |
| | | Unspecified acute lower respiratory infection # *J22* | |
| | | Respiratory failure, not elsewhere classified # *J96* | Respiratory failure # *518.81, 518.83, 518.84* |
| Pneumonia hospitalization | Primary diagnosis | *J12 – J18* | *480 – 486* |

Abbreviations: COPD (chronic obstructive pulmonary disease), ICD (International statistical classification of disease and related health problems)

**Table E3.** Baseline characteristics of participants with COPD aged ≥40 years in the HUNT2 study (1995-1997) followed for up to 20.4 years.

| COPD severity | | FEV1 z-score (Mean±SD) | FVC z-score (Mean±SD) | FEV1/FVC z-score (Mean±SD) | FEV1/FVC (Mean±SD) (%) |
|---|---|---|---|---|---|
| All | | -2.2±1.0 | -0.7±1.2 | -2.6±0.7 | 0.55± 0.08 |
| ppFEV$_1$ quartiles | 1 | -0.9±0.7 | 0.5±0.8 | -2.1±0.4 | 0.6±0.04 |
| | 2 | -1.9±0.3 | -0.4±0.6 | -2.4±0.5 | 0.6±0.05 |
| | 3 | -2.5±0.3 | -1.0±0.7 | -2.7±0.6 | 0.5±0.06 |
| | 4 | -3.4±0.6 | -2.0±0.9 | -3.3±0.8 | 0.5±0.08 |
| GOLD grades | 1 | -0.7±0.6 | 0.8±0.8 | -2.1±0.3 | 0.6±0.04 |
| | 2 | -2.2±0.5 | -0.7±0.7 | -2.6±0.6 | 0.6±0.06 |
| | 3 | -3.4±0.5 | -2.0±0.8 | -3.3±0.7 | 0.5±0.08 |
| | 4 | -4.4±0.6 | -3.1±0.8 | -4.2±0.7 | 0.4±0.09 |
| ATS/ERS grades | 1 | -1.2±0.7 | 0.2±0.8 | 2.2±0.4 | 0.6±0.05 |
| | 2 | -2.4±0.4 | -0.9±0.7 | -2.7±0.6 | 0.5±0.06 |
| | 3 | -3.3±0.4 | -1.9±0.8 | -3.3±0.7 | 0.5±0.08 |
| | 4 | -4.2±0.5 | -2.8±0.8 | -4.0±0.8 | 0.4±0.08 |
| FEV$_1$ z-score quartiles | 1 | -0.9±0.6 | 0.5±0.8 | -2.1±0.3 | 0.6±0.05 |
| | 2 | -1.9±0.2 | -0.4±0.6 | -2.5±0.5 | 0.6±0.06 |
| | 3 | -2.5±0.2 | -1.1±0.6 | -2.7±0.6 | 0.5±0.07 |
| | 4 | -3.5±0.5 | -1.9±0.9 | -3.4±0.7 | 0.5±0.09 |
| FEV$_1$ z-score grades | 1 | -1.2±0.7 | 0.2±0.8 | -2.2±0.4 | 0.6±0.06 |
| | 2 | -2.4±0.3 | -0.9±0.6 | -2.7±0.6 | 0.5±0.07 |
| | 3 | -3.4±0.3 | -2.0±0.8 | -3.4±0.6 | 0.5±0.08 |
| | 4 | -4.4±0.4 | -2.7±1.1 | -4.1±0.8 | 0.4±0.11 |
| FEV$_1$.Ht$^{-2}$ quartiles | 1 | -1.1±0.8 | 0.3±1.0 | -2.2±0.4 | 0.6±0.04 |
| | 2 | -1.9±0.6 | -0.4±0.8 | -2.5±0.6 | 0.58±0.06 |
| | 3 | -2.4±0.6 | -0.9±0.8 | -2.7±0.6 | 0.5±0.06 |
| | 4 | -3.3±0.7 | -1.9±0.9 | -3.2±0.8 | 0.5±0.09 |
| FEV$_1$.Ht$^{-2}$ grades | 1 | -1.8±0.9 | -0.3±1.0 | -2.5±0.6 | 0.6±0.06 |
| | 2 | -2.9±0.6 | -1.5±0.8 | -2.9±0.7 | 0.5±0.07 |
| | 3 | -3.5±0.5 | -2.1±0.9 | -3.5±0.7 | 0.5±0.08 |
| | 4 | -4.1±0.7 | -2.9±0.9 | -3.9±0.9 | 0.4±0.10 |
| FEV$_1$.Ht$^{-3}$ quartiles | 1 | -1.2±0.8 | 0.3±0.9 | -2.2±0.4 | 0.6±0.04 |

| | | | | | |
|---|---|---|---|---|---|
| | 2 | -1.9±0.6 | -0.4±0.8 | -2.5±0.6 | 0.6±0.05 |
| | 3 | -2.4±0.6 | -0.9±0.8 | -2.7±0.6 | 0.5±0.06 |
| | 4 | -3.3±0.7 | -1.9±0.9 | -3.3±0.8 | 0.5±0.08 |
| $FEV_1$.Q quartiles | 1 | -1.2±0.8 | 0.3±1.0 | -2.1±0.4 | 0.6±0.04 |
| | 2 | -2.0±0.7 | -0.4±0.8 | -2.5±0.6 | 0.6±0.05 |
| | 3 | -2.4±0.7 | -0.8±0.8 | -2.8±0.7 | 0.5±0.06 |
| | 4 | -3.2±0.7 | -1.8±0.9 | -3.2±0.8 | 0.5±0.08 |

Abbreviations: COPD (chronic obstructive pulmonary disease), ATS/ERS (American Thoracic Society/European Respiratory Society), GOLD (global initiative for chronic obstructive lung disease), HUNT (Nord-Trøndelag Health Study), SD (standard deviation), $ppFEV_1$ (percent-predicted forced expiratory volume in first second based in GLI-2012 equation, $FEV_1$ z-score (forced expiratory volume in first second z-score based on GLI-2012 equation), FVC z-score (forced vital capacity z-score based on GLI-2012 equation), $FEV_1.Ht^{-2}$ ($FEV_1$ standardized by square of height in meters), $FEV_1.Ht^{-3}$ ($FEV_1$ standardized by cube of height in meters), $FEV_1Q$ ($FEV_1$ standardized by sex-specific lowest percentile (0.5L for men and 0.4L for women) of $FEV_1$ distribution).

**Figure E2.** Cumulative incidence curves of classifications of COPD severity for respiratory mortality among participants with COPD aged ≥40 years in the HUNT2 study (1995-1997) followed for up to 20.4 years.

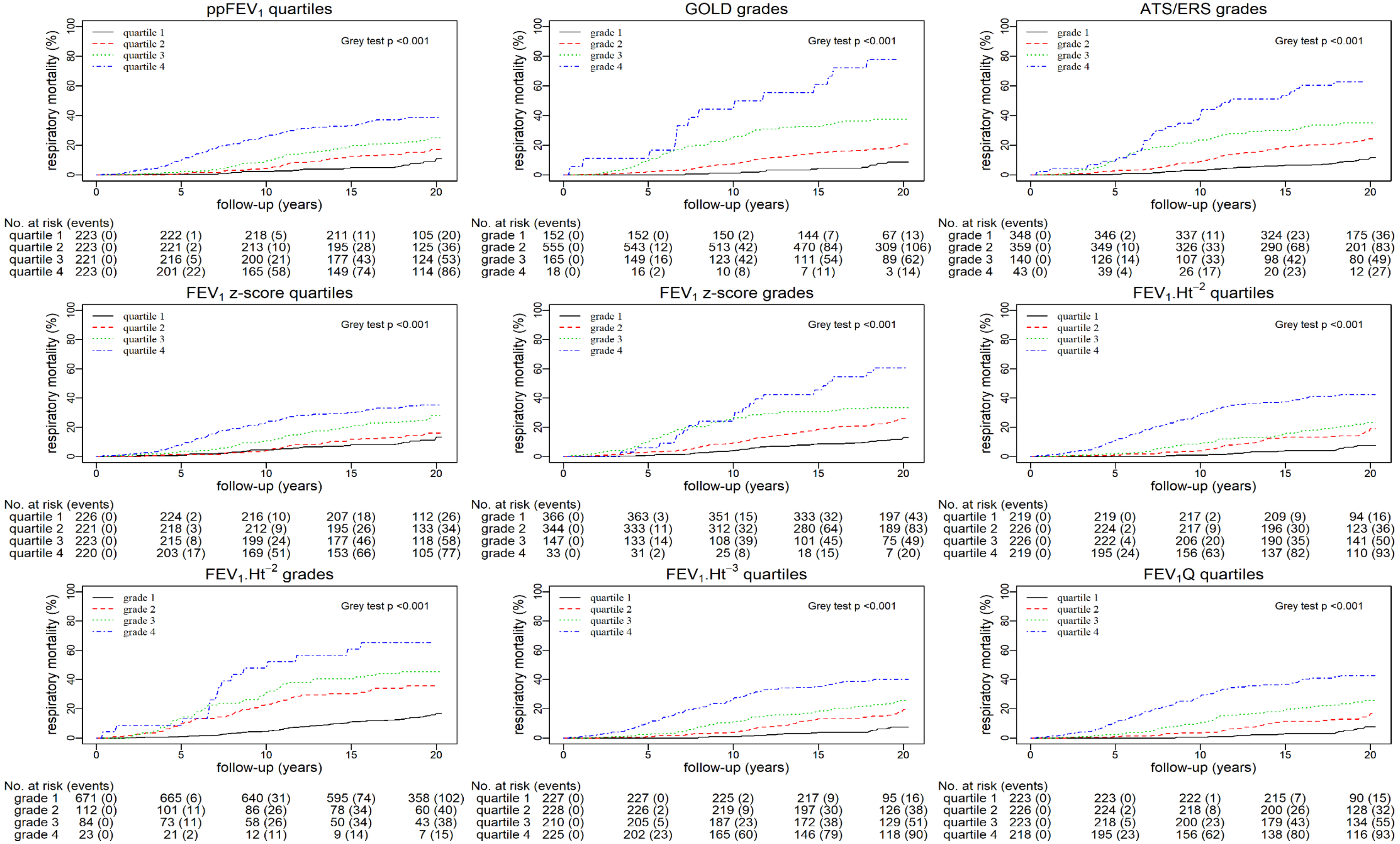

**Figure E3.** Cumulative incidence curves of classifications of COPD severity for cardiovascular mortality among participants with COPD aged ≥40 years in the HUNT2 study (1995-1997) followed for up to 20.4 years.

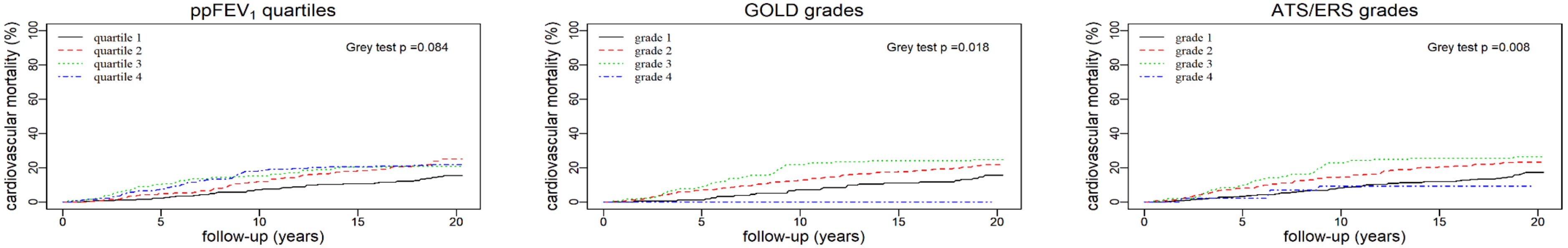


| No. at risk (events) | 0 | 5 | 10 | 15 | 20 |
|---|---|---|---|---|---|
| quartile 1 | 223 (0) | 218 (5) | 207 (16) | 198 (24) | 91 (34) |
| quartile 2 | 223 (0) | 212 (11) | 196 (27) | 182 (41) | 106 (55) |
| quartile 3 | 221 (0) | 198 (23) | 187 (34) | 175 (45) | 131 (46) |
| quartile 4 | 223 (0) | 206 (17) | 182 (41) | 177 (46) | 151 (49) |

| No. at risk (events) | 0 | 5 | 10 | 15 | 20 |
|---|---|---|---|---|---|
| grade 1 | 152 (0) | 150 (2) | 141 (11) | 134 (17) | 57 (23) |
| grade 2 | 555 (0) | 516 (39) | 484 (71) | 455 (99) | 295 (120) |
| grade 3 | 165 (0) | 150 (15) | 129 (36) | 125 (40) | 110 (41) |
| grade 4 | 18 (0) | 18 (0) | 18 (0) | 18 (0) | 17 (0) |

| No. at risk (events) | 0 | 5 | 10 | 15 | 20 |
|---|---|---|---|---|---|
| grade 1 | 348 (0) | 336 (12) | 319 (29) | 305 (42) | 152 (59) |
| grade 2 | 359 (0) | 330 (29) | 306 (53) | 284 (74) | 200 (84) |
| grade 3 | 140 (0) | 126 (14) | 108 (32) | 104 (36) | 92 (37) |
| grade 4 | 43 (0) | 39 (1) | 39 (4) | 39 (4) | 35 (4) |

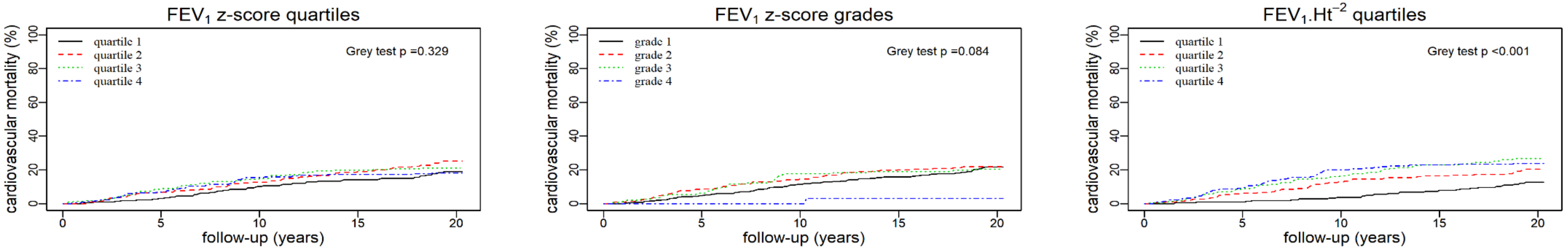


| No. at risk (events) | 0 | 5 | 10 | 15 | 20 |
|---|---|---|---|---|---|
| quartile 1 | 226 (0) | 219 (7) | 203 (23) | 193 (32) | 96 (42) |
| quartile 2 | 221 (0) | 207 (14) | 193 (28) | 179 (42) | 112 (55) |
| quartile 3 | 223 (0) | 203 (20) | 190 (33) | 179 (44) | 129 (47) |
| quartile 4 | 220 (0) | 205 (15) | 186 (34) | 181 (38) | 142 (40) |

| No. at risk (events) | 0 | 5 | 10 | 15 | 20 |
|---|---|---|---|---|---|
| grade 1 | 366 (0) | 349 (17) | 324 (42) | 307 (58) | 162 (78) |
| grade 2 | 344 (0) | 314 (30) | 294 (50) | 275 (69) | 197 (75) |
| grade 3 | 147 (0) | 138 (9) | 121 (26) | 118 (28) | 94 (30) |
| grade 4 | 33 (0) | 33 (0) | 33 (0) | 32 (1) | 26 (1) |

| No. at risk (events) | 0 | 5 | 10 | 15 | 20 |
|---|---|---|---|---|---|
| quartile 1 | 219 (0) | 217 (2) | 211 (8) | 201 (17) | 83 (27) |
| quartile 2 | 226 (0) | 212 (14) | 197 (29) | 189 (37) | 114 (45) |
| quartile 3 | 226 (0) | 207 (19) | 189 (37) | 173 (52) | 131 (60) |
| quartile 4 | 219 (0) | 198 (21) | 175 (44) | 169 (50) | 151 (52) |

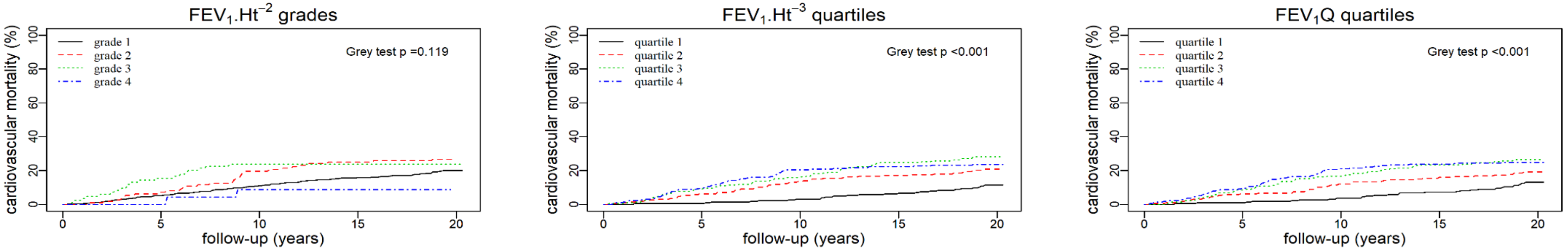


| No. at risk (events) | 0 | 5 | 10 | 15 | 20 |
|---|---|---|---|---|---|
| grade 1 | 671 (0) | 636 (35) | 597 (74) | 563 (106) | 328 (132) |
| grade 2 | 112 (0) | 104 (8) | 90 (22) | 84 (28) | 70 (30) |
| grade 3 | 84 (0) | 71 (13) | 64 (20) | 64 (20) | 61 (20) |
| grade 4 | 23 (0) | 23 (0) | 21 (2) | 21 (2) | 20 (2) |

| No. at risk (events) | 0 | 5 | 10 | 15 | 20 |
|---|---|---|---|---|---|
| quartile 1 | 227 (0) | 226 (1) | 220 (7) | 211 (15) | 86 (25) |
| quartile 2 | 228 (0) | 214 (14) | 197 (31) | 188 (39) | 117 (47) |
| quartile 3 | 210 (0) | 191 (19) | 176 (34) | 158 (52) | 121 (59) |
| quartile 4 | 225 (0) | 203 (22) | 179 (46) | 175 (50) | 155 (53) |

| No. at risk (events) | 0 | 5 | 10 | 15 | 20 |
|---|---|---|---|---|---|
| quartile 1 | 223 (0) | 221 (2) | 215 (8) | 206 (16) | 77 (28) |
| quartile 2 | 226 (0) | 212 (14) | 199 (27) | 190 (36) | 117 (43) |
| quartile 3 | 223 (0) | 204 (19) | 186 (37) | 170 (52) | 130 (59) |
| quartile 4 | 218 (0) | 197 (21) | 172 (46) | 166 (52) | 155 (54) |

**Figure E4.** Cumulative incidence curves of classifications of COPD severity for COPD hospitalization among participants with COPD aged ≥40 years in the HUNT2 study (1995-1997) followed for up to 20.4 years.

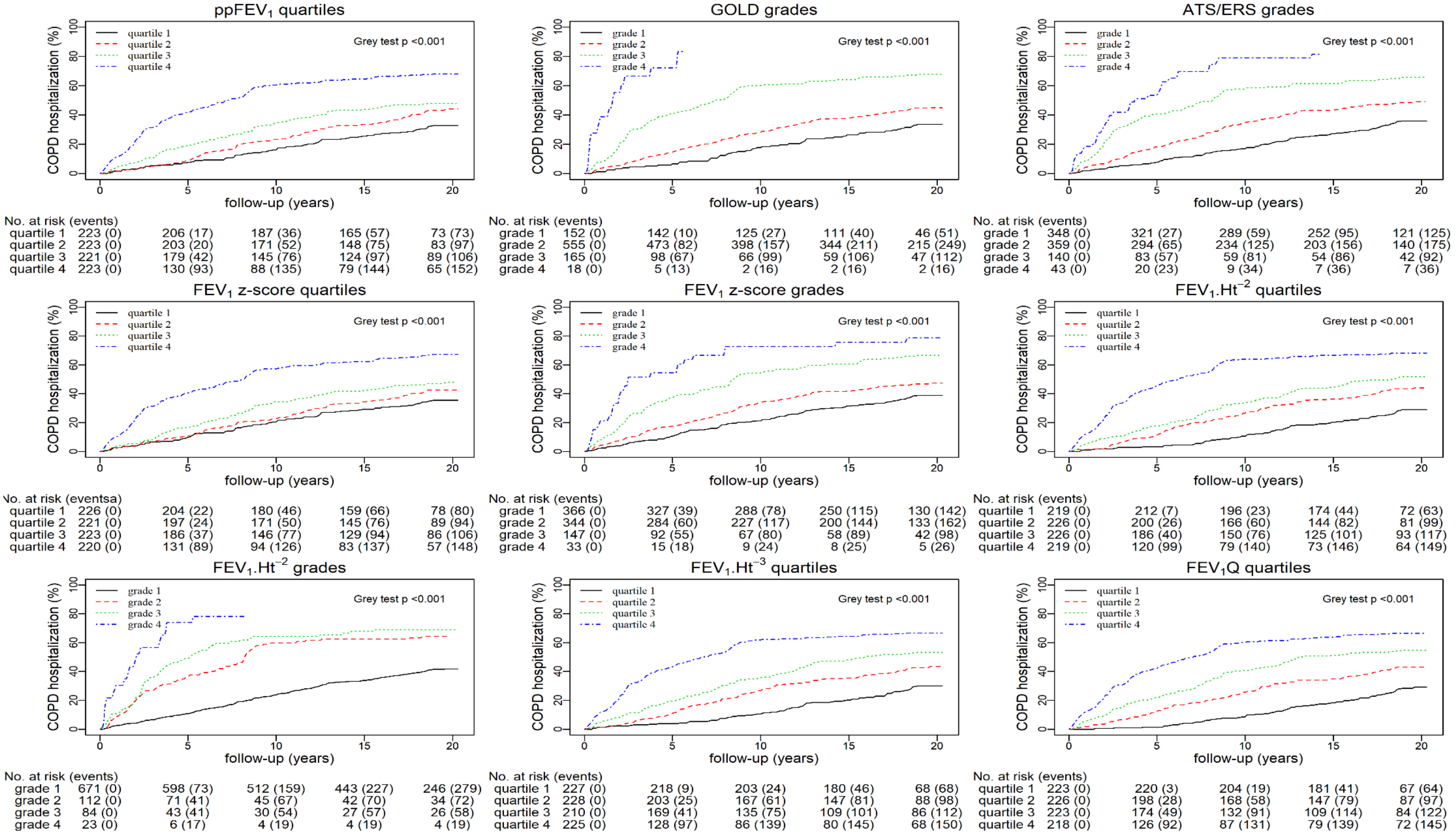

**Figure E5.** Cumulative incidence curves of classifications of COPD severity for pneumonia hospitalization among participants with COPD aged ≥40 years in the HUNT2 study (1995-1997) followed for up to 20.4 years.

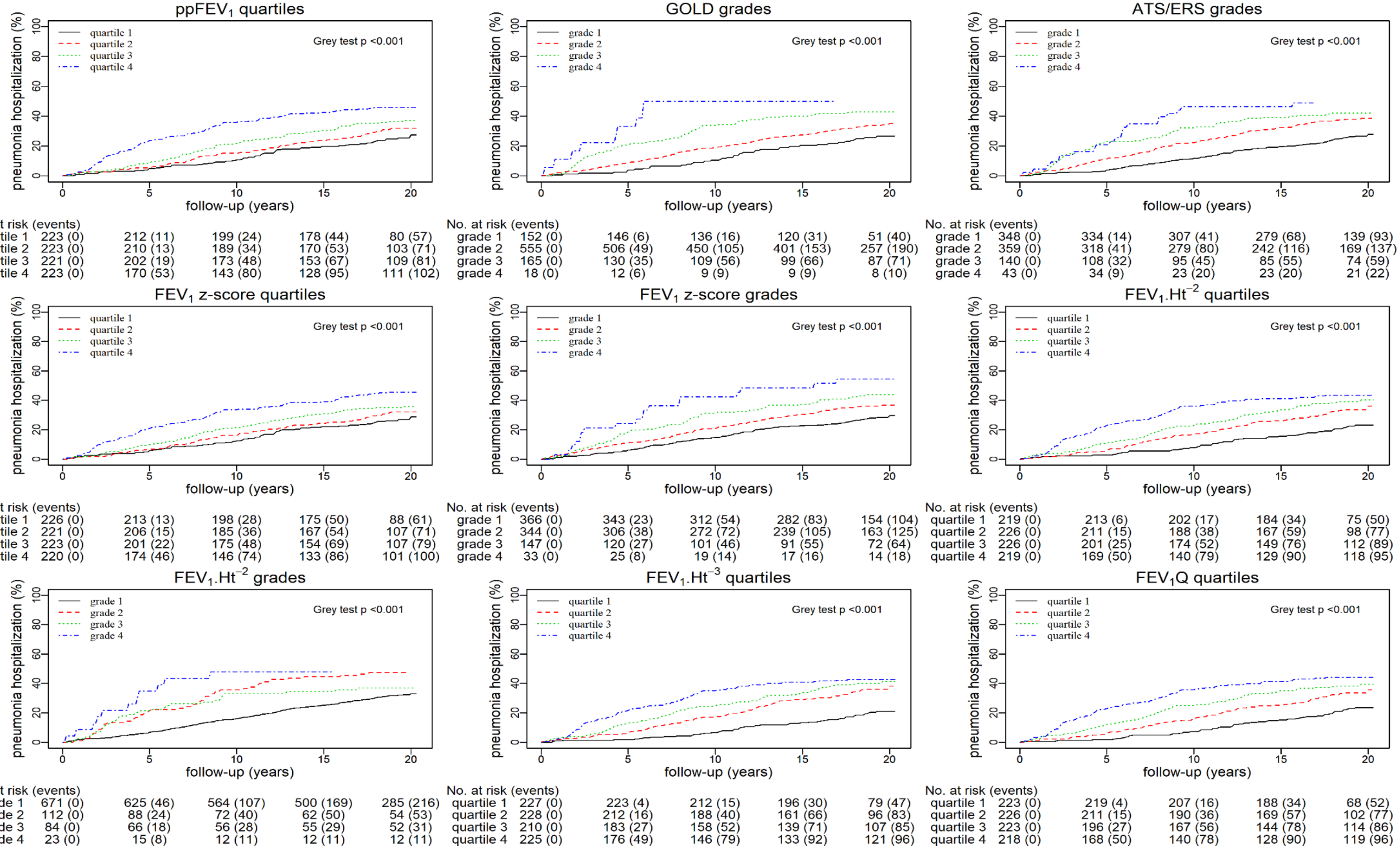


**ppFEV$_1$ quartiles** — No. at risk (events)

| | 0 | 5 | 10 | 15 | 20 |
|---|---|---|---|---|---|
| quartile 1 | 223 (0) | 212 (11) | 199 (24) | 178 (44) | 80 (57) |
| quartile 2 | 223 (0) | 210 (13) | 189 (34) | 170 (53) | 103 (71) |
| quartile 3 | 221 (0) | 202 (19) | 173 (48) | 153 (67) | 109 (81) |
| quartile 4 | 223 (0) | 170 (53) | 143 (80) | 128 (95) | 111 (102) |

**GOLD grades** — No. at risk (events)

| | 0 | 5 | 10 | 15 | 20 |
|---|---|---|---|---|---|
| grade 1 | 152 (0) | 146 (6) | 136 (16) | 120 (31) | 51 (40) |
| grade 2 | 555 (0) | 506 (49) | 450 (105) | 401 (153) | 257 (190) |
| grade 3 | 165 (0) | 130 (35) | 109 (56) | 99 (66) | 87 (71) |
| grade 4 | 18 (0) | 12 (6) | 9 (9) | 9 (9) | 8 (10) |

**ATS/ERS grades** — No. at risk (events)

| | 0 | 5 | 10 | 15 | 20 |
|---|---|---|---|---|---|
| grade 1 | 348 (0) | 334 (14) | 307 (41) | 279 (68) | 139 (93) |
| grade 2 | 359 (0) | 318 (41) | 279 (80) | 242 (116) | 169 (137) |
| grade 3 | 140 (0) | 108 (32) | 95 (45) | 85 (55) | 74 (59) |
| grade 4 | 43 (0) | 34 (9) | 23 (20) | 23 (20) | 21 (22) |

**FEV$_1$ z-score quartiles** — No. at risk (events)

| | 0 | 5 | 10 | 15 | 20 |
|---|---|---|---|---|---|
| quartile 1 | 226 (0) | 213 (13) | 198 (28) | 175 (50) | 88 (61) |
| quartile 2 | 221 (0) | 206 (15) | 185 (36) | 167 (54) | 107 (71) |
| quartile 3 | 223 (0) | 201 (22) | 175 (48) | 154 (69) | 107 (79) |
| quartile 4 | 220 (0) | 174 (46) | 146 (74) | 133 (86) | 101 (100) |

**FEV$_1$ z-score grades** — No. at risk (events)

| | 0 | 5 | 10 | 15 | 20 |
|---|---|---|---|---|---|
| grade 1 | 366 (0) | 343 (23) | 312 (54) | 282 (83) | 154 (104) |
| grade 2 | 344 (0) | 306 (38) | 272 (72) | 239 (105) | 163 (125) |
| grade 3 | 147 (0) | 120 (27) | 101 (46) | 91 (55) | 72 (64) |
| grade 4 | 33 (0) | 25 (8) | 19 (14) | 17 (16) | 14 (18) |

**FEV$_1$.Ht$^{-2}$ quartiles** — No. at risk (events)

| | 0 | 5 | 10 | 15 | 20 |
|---|---|---|---|---|---|
| quartile 1 | 219 (0) | 213 (6) | 202 (17) | 184 (34) | 75 (50) |
| quartile 2 | 226 (0) | 211 (15) | 188 (38) | 167 (59) | 98 (77) |
| quartile 3 | 226 (0) | 201 (25) | 174 (52) | 149 (76) | 112 (89) |
| quartile 4 | 219 (0) | 169 (50) | 140 (79) | 129 (90) | 118 (95) |

**FEV$_1$.Ht$^{-2}$ grades** — No. at risk (events)

| | 0 | 5 | 10 | 15 | 20 |
|---|---|---|---|---|---|
| grade 1 | 671 (0) | 625 (46) | 564 (107) | 500 (169) | 285 (216) |
| grade 2 | 112 (0) | 88 (24) | 72 (40) | 62 (50) | 54 (53) |
| grade 3 | 84 (0) | 66 (18) | 56 (28) | 55 (29) | 52 (31) |
| grade 4 | 23 (0) | 15 (8) | 12 (11) | 12 (11) | 12 (11) |

**FEV$_1$.Ht$^{-3}$ quartiles** — No. at risk (events)

| | 0 | 5 | 10 | 15 | 20 |
|---|---|---|---|---|---|
| quartile 1 | 227 (0) | 223 (4) | 212 (15) | 196 (30) | 79 (47) |
| quartile 2 | 228 (0) | 212 (16) | 188 (40) | 161 (66) | 96 (83) |
| quartile 3 | 210 (0) | 183 (27) | 158 (52) | 139 (71) | 107 (85) |
| quartile 4 | 225 (0) | 176 (49) | 146 (79) | 133 (92) | 121 (96) |

**FEV$_1$Q quartiles** — No. at risk (events)

| | 0 | 5 | 10 | 15 | 20 |
|---|---|---|---|---|---|
| quartile 1 | 223 (0) | 219 (4) | 207 (16) | 188 (34) | 68 (52) |
| quartile 2 | 226 (0) | 211 (15) | 190 (36) | 169 (57) | 102 (77) |
| quartile 3 | 223 (0) | 196 (27) | 167 (56) | 144 (78) | 114 (86) |
| quartile 4 | 218 (0) | 168 (50) | 140 (78) | 128 (90) | 119 (96) |

**Figure E6.** Adjusted hazard ratios[#] for different expressions of $FEV_1$ and their respective methods of classification of COPD severity for all-cause mortality, respiratory mortality, cardiovascular mortality, COPD hospitalization, and pneumonia hospitalization among participants with COPD aged ≥40 years in the HUNT2 study (1995-1997) followed for up to 20.4 years.

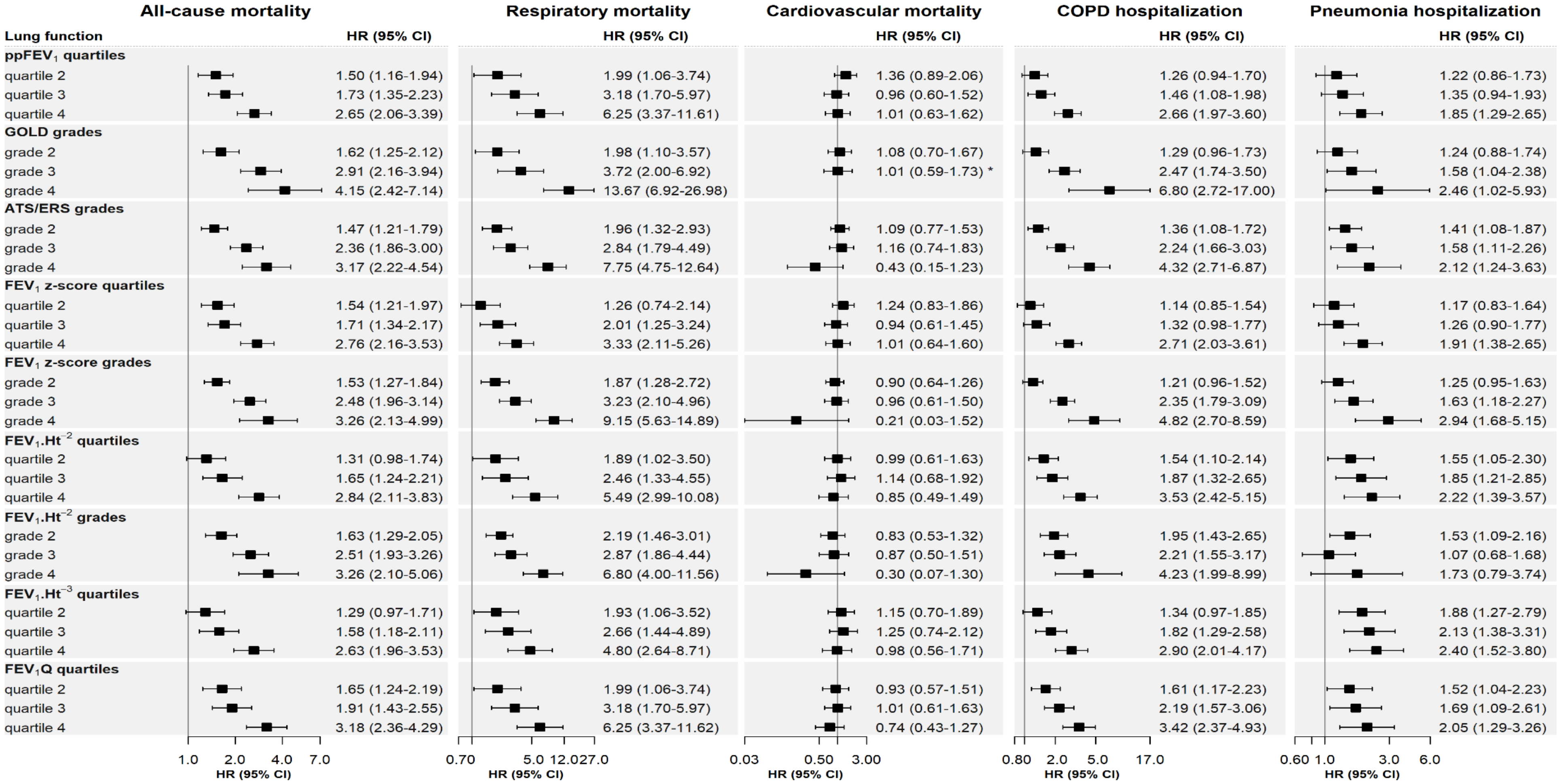


Abbreviations: COPD (chronic obstructive pulmonary disease), ATS/ERS (American Thoracic Society/European Respiratory Society), GOLD (global initiative for chronic obstructive lung disease), HUNT (Nord-Trøndelag Health Study), CI (confidence interval), HR (hazard ratios), $ppFEV_1$ (percent-predicted forced expiratory volume in first second based in GLI-2012 equation, $FEV_1$ z-score (forced expiratory volume in first second z-score based

on GLI-2012 equation), $FEV_1.Ht^{-2}$ ($FEV_1$ standardized by square of height in meters), $FEV_1.Ht^{-3}$ ($FEV_1$ standardized by cube of height in meters), $FEV_1Q$ ($FEV_1$ standardized by sex-specific lowest percentile (0.5L for men and 0.4L for women) of $FEV_1$ distribution).
[#]- adjusted for age, sex, smoking, body mass index, education, *- GOLD grades 3-4 were combined due to zero cases in grade 4.

**Figure E7.** Multivariate regression trees for $FEV_1.Ht^{-2}$, $FEV_1.Ht^{-3}$, and $FEV_1Q$ in predicting multiple outcomes respiratory mortality, cardiovascular mortality, other cause-related mortality, and COPD hospitalization among participants with COPD aged ≥40 years in the HUNT2 study (1995-1997) followed for up to 20.4 years.

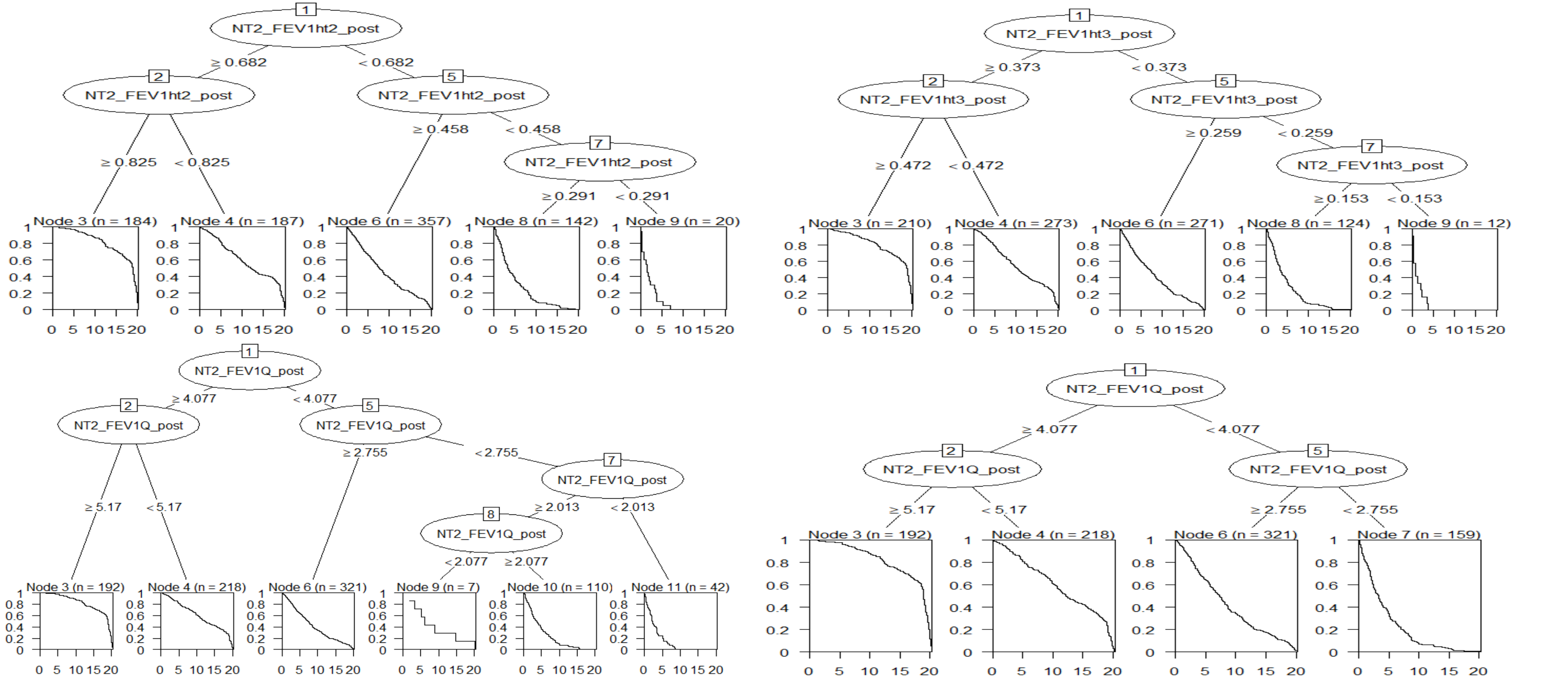


Abbreviations: COPD (chronic obstructive pulmonary disease), HUNT (Nord-Trøndelag Health Study), $FEV_1.Ht^{-2}$ (forced expiratory volume in first second ($FEV_1$) standardized by square of height in meters), $FEV_1.Ht^{-3}$ ($FEV_1$ standardized by cube of height in meters), $FEV_1Q$ ($FEV_1$ standardized by sex-specific lowest percentile (0.5L for men and 0.4L for women) of $FEV_1$ distribution).

### UK Biobank

**Text E1.** Study population in UK Biobank.

The UK Biobank is a prospective cohort study, which was conducted during 2006 to 2010 [1]. Among 9 million invited, 344 792 people aged 40-69 years were recruited to the UK Biobank from 22 recruitment centres throughout UK [1,2]. The participants with information on lung function were 242 459. There were 6495 participants with COPD when we defined COPD as having pre-bronchodilator $FEV_1/FVC<0.70$ (fixed-ratio criteria [3]) and pre-bronchodilator $FEV_1/FVC$ z-scores < -1.645 (lower limit of normal (LLN) criteria [4,5]) and [respiratory symptoms (wheezing or dyspnoea) and/or self-reported doctor-diagnosed COPD]. We excluded the participants who reported of smoking or have used an inhaler within an hour prior to spirometry test.

The prebronchodilator lung function test was performed using a handheld pneumotachograph spirometer (Pneumotrac 6800) [2]. Quality assurance of spirometric measurements is described in detail elsewhere [2,6].

**Figure E8.** Distribution of participants with COPD, all-cause mortality, respiratory mortality, and cardiovascular mortality in categories of GOLD grades and $FEV_1Q$ grades among participants with COPD aged ≥40 years in the UK Biobank (2006-2010) followed for 5 years.

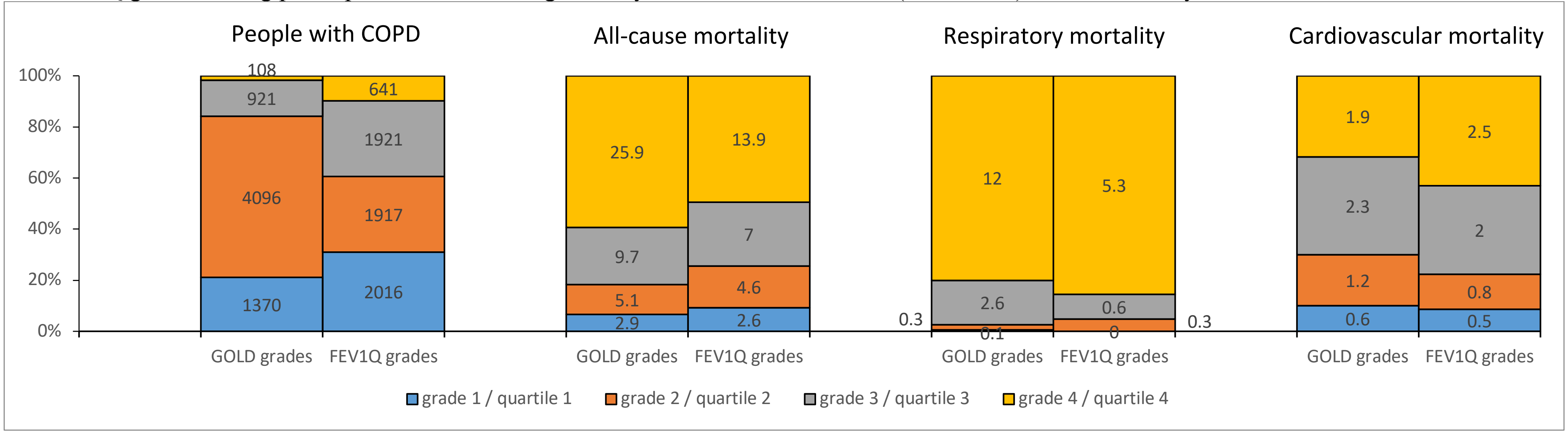

Abbreviations: COPD (chronic obstructive pulmonary disease), GOLD (global initiative for chronic obstructive lung disease), $FEV_1Q$ ($FEV_1$ standardized by sex-specific lowest percentile (0.5L for men and 0.4L for women) of $FEV_1$ distribution).
Note- The optimal cut-offs of $FEV_1Q$ ($FEV_1Q$ grades) generated in HUNT were tested in UK Biobank.

**Figure E9.** The AUCs for different expressions of $FEV_1$ and their respective methods of classification of COPD severity for all-cause mortality, respiratory mortality, and cardiovascular mortality among participants with COPD aged ≥40 years in the UK Biobank (2006-2010) followed for 5 years.

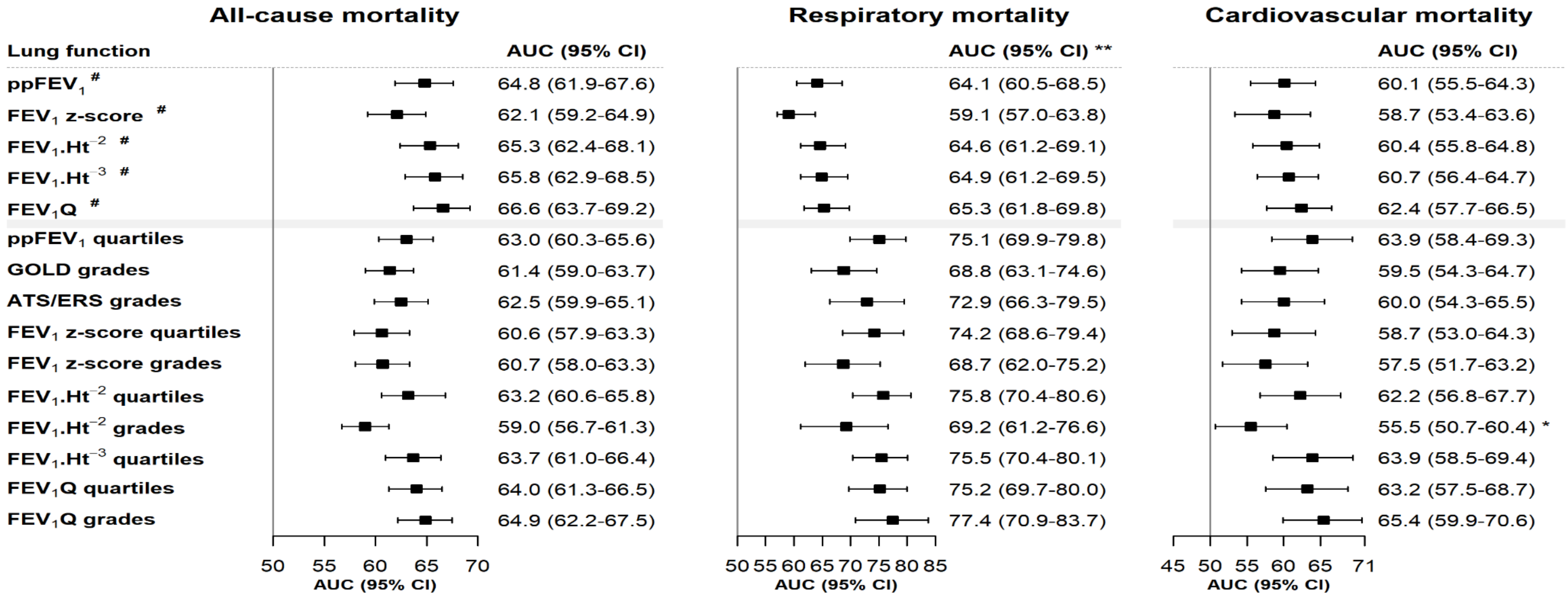


Abbreviations: COPD (chronic obstructive pulmonary disease), ATS/ERS (American Thoracic Society/European Respiratory Society), GOLD (global initiative for chronic obstructive lung disease), CI (confidence interval), AUC (area under receiver operating characteristics curves), $ppFEV_1$ (percent-predicted forced expiratory volume in first second based in GLI-2012 equation, $FEV_1$ z-score (forced expiratory volume in first second z-score based on GLI-2012 equation), $FEV_1.Ht^{-2}$ ($FEV_1$ standardized by square of height in meters), $FEV_1.Ht^{-3}$ ($FEV_1$ standardized by cube of height in meters), $FEV_1Q$ ($FEV_1$ standardized by sex-specific lowest percentile (0.5L for men and 0.4L for women) of $FEV_1$ distribution).
[#]- continuous variables, *- $FEV_1.Ht^{-2}$ grades 3-4 were combined due to zero cases in grade 4, **- grades/quartiles 2-4 were analysed due to zero cases in grade/quartile 1 of $FEV_1Q$ quartiles and $FEV_1Q$ grades. **- Note: similar differences in AUCs were observed when grade/quartile 1-2 were combined for respiratory mortality. Note- The optimal cut-offs of $FEV_1Q$ ($FEV_1Q$ grades) generated in HUNT were tested in UK Biobank.

#### References:

1. UK Biobank. 2006-2010, https://www.ukbiobank.ac.uk/, Accessed 01 February 2019.
2. Gupta RP, Strachan DP. Ventilatory function as a predictor of mortality in lifelong non-smokers: evidence from large British cohort studies. *BMJ Open* 2017; **7**(7).
3. Global Initiative for Chronic Obstructive Lung Disease. Global strategy for the diagnosis, management, and prevention of chronic obstructive pulmonary diesease, 2019. [Accessed 01 January 2020] Available from URL: http://goldcopd.org/.
4. Pellegrino R, Viegi G, Brusasco V, et al. Interpretative strategies for lung function tests. *The European respiratory journal* 2005; **26**(5): 948-68.
5. Quanjer PH, Stanojevic S, Cole TJ, et al. MULTI-ETHNIC REFERENCE VALUES FOR SPIROMETRY FOR THE 3–95 YEAR AGE RANGE: THE GLOBAL LUNG FUNCTION 2012 EQUATIONS: Report of the Global Lung Function Initiative (GLI), ERS Task Force to establish improved Lung Function Reference Values. *The European respiratory journal* 2012; **40**(6): 1324-43.
6. De Matteis S, Jarvis D, Hutchings S, et al. Occupations associated with COPD risk in the large population-based UK Biobank cohort study. *Occup Environ Med* 2016; **73**(6): 378-84.
